# CDC (Cindy and David’s Conversations) Game: Advising President to Survive Pandemic

**DOI:** 10.1101/2022.03.14.22272381

**Authors:** Zhanshan (Sam) Ma, Liexun Yang

## Abstract

Ongoing debates on anti-COVID19 policies have been focused on coexistence *vs*. zero-out strategies, which can be simplified as “always open (AO)” *vs*. “always closed (AC).” We postulate that, the middle ground between the two extremes, dubbed LOHC (low-risk open and high-risk closed), is likely more favorable, precluding obviously irrational HOLC (high-open-low-closed). From a meta-strategy perspective, these four policies cover the full spectrum of anti-pandemic policies. We argue that, among numerous factors influencing strategic policy-making, the competence of advisory body such as CDC chief-scientist (*say*, Cindy) and politics in decision-making body such as president (David), and their cooperation/communication can be critical. Here we investigate anti-pandemic policy-making by harnessing the power of evolutionary game theory in modeling competition/cooperation/communication (three critical processes underlying biological and social evolutions). Specifically, we apply the Sir Philip Sydney (SPS) game, a 4×4 signaler-responder evolutionary game with 16 strategic interactions, which was devised to investigate the reliability of communication that can modulate competition and cooperation, to capture rich idiosyncrasies surrounding today’s anti-pandemic policies. By emulating the reality of anti-pandemic policies today, the study aims to identify possible cognitive gaps and traps. The extended SPS, dubbed CDC (Cindy and David’s Conversations) game, offers a powerful cognitive model for investigating the coexistence/zero-out dichotomy and possible alternatives. The rigorous analytic solutions and extensive simulations suggest a take-home message—keep it persistently simple and rational: while apparently preferred middle-ground LOHC seems to be small-probability (∼0.05) event counter-intuitively, the AO and AC policies appears to be large-probability (∼0.41-0.53) events.

**Lay Summary:** Ongoing debates on anti-COVID19 policies have been focused on coexistence-with *vs*. zero-out (virus) strategies, which can be simplified as “always open (AO)” *vs*. “always closed (AC).” We postulate that middle ground, dubbed LOHC (low-risk-open and high-risk-closed), is likely more favorable, precluding obviously irrational HOLC (high-risk-open and low-risk-closed). From a meta-strategy perspective, these four policies cover the full spectrum of anti-pandemic policies. By emulating the reality of anti-pandemic policies today, the study aims to identify possible cognitive gaps and traps by harnessing the power of evolutionary game-theoretic analysis and simulations, which suggest that (*i*) AO and AC seems to be “high-probability” events (∼0.41-0.53); (*ii*) counter-intuitively, the middle ground—LOHC—seems to be small-probability event (∼0.05), possibly due to its unduly complexity, mirroring its wide-range failures in practice.

## Background—Cindy and David’s Conversations

*“During the Covid-19 response, the greatest challenge to public health in more than 100 years*, ***science*** *must guide public health decision making”* Sonja Rasmussen & Denise Jamieson (2020)

“[**Science**] *can inform policy, but it can’t dictate how to weigh the moral and political nature of policy makers’ decisions*.” Nason Maani & Sandro Galea (2021)

“*Human behavior evolves as our knowledge increases, but we are all subject to our own ways of construing this knowledge. Because of the pervasive influence of social media, new knowledge is often overwhelmed by* ***misinformation*** *that further confuses us and provides easy access to conspiracy theories and alternative facts*.” Nicholas Dirks (2021)

“*Another recurring theme in historical analyses of epidemics is that medical and public health interventions* ***often fail*** *to live up to their promise.”* David Jones (2020)

The COVID-19 pandemic has been pushing the debates on anti-pandemic policy-making to an unprecedented contentious level both nationally and internationally. Perhaps the only certainty in the short term would be no consensus due to the complex strategic interactions among scientific, socioeconomic and international political factors. Arguably, the focus of these strategic interactions can be abstracted as an advisory relationship between a country’s advisory body (*e.g*., CDC chief scientist) and state head (*e.g*., President). In an idealized world, the chief is supposed to fully grasp the pandemic status and capable to provide sound scientific recommendations to president in time, and the president is supposed to make decisions that benefits the country, if not the world, most. In reality, either chief or president may be faulty in understanding the state-of-the-pandemic and their communication channel may be “noisy”, which may lead to unexpected and/or undesirable consequences. “Faulty” can be misjudgment of pandemic risk, and “noisy” communication can be misinformation or miscommunication; fault and noise can be associated with either chief, president or both.

In the present study, we formulate the CDC-chief *vs*. president relationship as an evolutionary game to facilitate the analysis of their strategic interactions using a fictitious pair of players (Cindy as CDC chief, who represent the expertise authority of medical science, *vs*. David as President, the head of the decision-making body in general), and the CDC (Cindy and David’s Conversations) game is apparently inspired by the classic Sir Philip Sydney (SPS) game and its extensions (Maynard-Smith 1991, Maynard-Smith & Harper 2003, Bergstrom & Lachmann 1997, 1998, Huttegger & Zollman 2010, Ma 2009, Ma *et al*. 2010, Ma & Krings 2011, Whitmeyer 2020, Ma & Zhang 2021). We further take inspirations from classic ecological theories of metapopulation dynamics (Levins 1969, Citron *et al*. 2021) and Allee effects (Allee 1932, Hilker *et al*. 2009), in which the worldwide pandemic of COVID-19 can be abstracted as a dynamic system of *metapopulation* (of infected populations) consisting of approximately 200 local (country or regional) *populations*. The local populations are connected through migration/dispersal (such as international travels and trade logistics) and collectively form the global metapopulation, that is, *metapopulaton* is a *population* of *local populations*. According to metapopulation and Allee effects theories, local extinctions (zero-outs) can be as common as local outbreaks. Such phenomena may not only explain the highly heterogeneous distribution patterns of local outbreaks of the COVID-19 pandemic, but also shed lights on the intricacies of various anti-pandemic policies.

The CDC game is abstracted as an asymmetric four-by-four strategic-form game, 16 strategic interactions (Table 1, Fig 1). The justifications why we adopted evolutionary game modeling, specifically the SPS game, have high significances to do with the five unique, fundamental characteristics of the SPS evolutionary game, as exposed below. Before presenting the five characteristics, we argue for a general strategic principle, which maintains that three fundamental processes or forces—competition, cooperation and communication (or 3C)—drive the biological (social) evolutions, including the policy-making for anti-pandemic. We do not argue that both biological (organismal) evolution and social evolution (development) are the same, and we only argue that the tools to study organismal evolution can be harnessed to study questions in social evolution because the 3C processes (forces) are major driving forces for their evolution (development) in both systems. Darwin’s evolutionary theory focused on competition and struggling for living, but largely ignored cooperation, not to mention communication that we argue is able to modulate competition and cooperation. In the 1960s, the critical significance of *cooperation* in the form of kin selection was introduced to supplement Darwin’s evolutionary theory (Hamilton 1964). Since then, five major forms of cooperation have been identified and most notably (Axelrod & Hamilton 1981, Nowak 2006, Nowak & Sigmund 2007), Prisoner’s dilemma was introduced and recognized as one of the most powerful models for modeling cooperation in evolutionary biology and social-evolution studies (Aumann 1959, Amadae 2016, Tanimoto 2015, 2021). The handicap principle first proposed in the 1990s filled the gap left by the evolutionary theory (Zahavi 1975, 1997, Grafen 1990, Maynard-Smith 1990), and its theoretical incarnation spawned the invention of Sir Philip Sydney (SPS) game (Maynard-Smith 1991, Maynard-Smith & Harper 2003, Huttegger & Zollman 2010, Brown 2016, Cooper *et al*. 2018, Whitmeyer 2020, Ma & Zhang 2021, Zahavi 1975, 1997). In fact, evolutionary game theory (including SPS game model) was an integration of game theory and evolutionary theory (Maynard-Smith & Price 1973, Grafen 1990, Amadae 2016, Ma 2009, Ma *et al*. 2010, Ma & Krings 2011), and the integration turned evolutionary game theory into one of the most active branches of game theory.

**Table 1.** From Sir Philip Sidney (SPS) game to Cindy and David’s Conversations (CDC) game

| The Sir Philip Sydney (SPS) Game and Its Extensions<br>(Maynard-Smith 1992, Huttegger & Zollman 2010, Ma & Krings 2011, Whitmeyer 2020, Ma & Zhang 2021) |  |  |  |  |  |
| --- | --- | --- | --- | --- | --- |
| Sender (Signaler) Strategies |  | Responder (Donor) Strategies |  |  |  |
| S<sub>1</sub>: Signal only if healthy |  | R<sub>1</sub>: Donate only if no signal |  |  |  |
| S<sub>2</sub>: Signal only if needy |  | R<sub>2</sub>: Donate only if signal |  |  |  |
| S<sub>3</sub>: Never signal |  | R<sub>3</sub>: Never donate |  |  |  |
| S<sub>4</sub>: Always signal |  | R<sub>4</sub>: Always donate |  |  |  |
| Cindy and David's Conversations (CDC) game |  |  |  |  |  |
| Cindy's Strategies (Advices) |  | David's Strategies (Decisions) |  |  |  |
| C<sub>1</sub>: Open only if low-risk |  | D<sub>1</sub>: Closed only if Cindy advises closed |  |  |  |
| C<sub>2</sub>: Open only if high-risk |  | D<sub>2</sub>: Closed only if Cindy advises open |  |  |  |
| C<sub>3</sub>: Always closed |  | D<sub>3</sub>: Always open |  |  |  |
| C<sub>4</sub>: Always open |  | D<sub>4</sub>: Always closed |  |  |  |
| For examples:<br>C<sub>1</sub> is intuitively equivalent to:<br>*Open* = When low-risk with probability (1−*m*);<br>*Closed* = When high-risk with probability (*m*). |  | For examples:<br>D<sub>1</sub> is intuitively equivalent to: *Closed*=Following<br>Cindy's advice of closed with potential loss of *d*<br>(such as dissatisfaction of public); *Open*=following<br>Cindy's advice of open without potential loss of *d*. |  |  |  |
| The 4x4 combinatorial asymmetric strategies of the discrete CDC game<br>with the letters (A-P) for possible strategy pairs as shown in Fig 1 |  |  |  |  |  |
| Responder's strategies<br>(David's decisions)<br>Sender strategies<br>(Cindy's Advices) |  | D<sub>1</sub><br>Closed only if<br>Cindy advises<br>Closed | D<sub>2</sub><br>Closed only if<br>Cindy advises<br>Open | D<sub>3</sub><br>Always<br>Open | D<sub>4</sub><br>Always<br>Closed |
| C<sub>1</sub>: Open only if low-risk |  | A | B | C | D |
| C<sub>2</sub>: Open only if high-risk |  | E | F | G | H |
| C<sub>3</sub>: Always closed |  | I | J | K | L |
| C<sub>4</sub>: Always open |  | M | N | O | P |
| Four Basic Anti-Pandemic Policies, which should be the output from the previous CDC game<br>model (see Table 2 for the corresponding CDC equilibriums) |  |  |  |  |  |
| Policy # | Policy Description | Possible CDC equilibriums likely to output<br>the policies |  |  | Example |
| 1 | Low (Risk) Open & High (Risk)<br>Closed = **LOHC** | Strategy A, Nash Equilibrium; Strategy F,<br>ESS (evolutionarily stable strategies);<br>Polymorphic Hybrid Equilibrium (PHE) |  |  | Near<br>universally<br>adopted |
| 2 | High (Risk) Open & Low (Risk)<br>Closed = **HOLC** | No equilibriums possible; but B & E as<br>non-equilibrium strategies are possible. |  |  | Unlikely to<br>sustain |
| 3 | Always Open = **AO** | Mixed Strategy J & K, Pooling<br>Equilibrium; PHE |  |  | Herd<br>immunity |
| 4 | Always Closed = **AC** | Mixed Strategy I & L; Strategy L,<br>Pooling Equilibrium; PHE |  |  | Zero-out |

**Fig 1.**
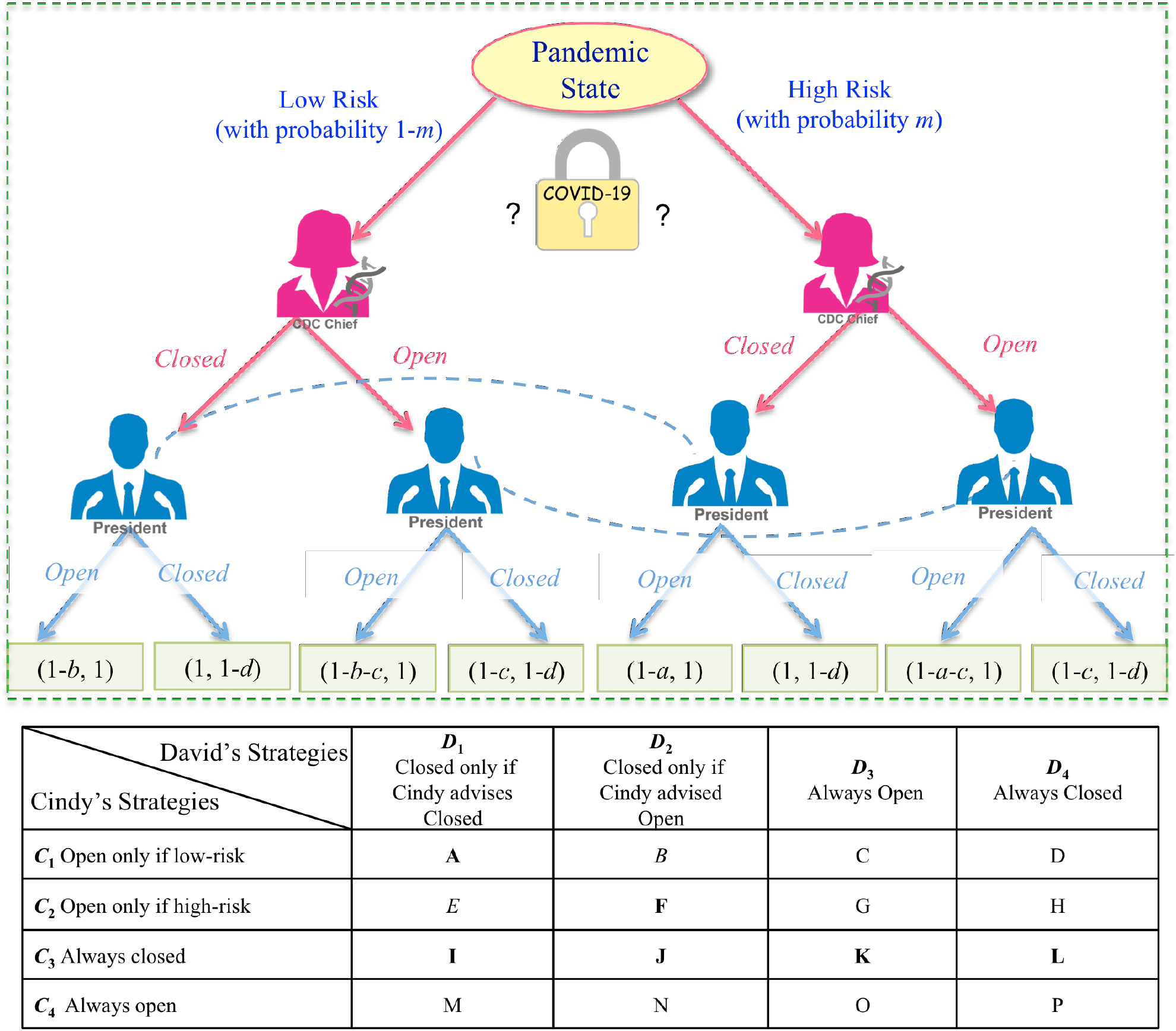
A decision-tree of CDC (Cindy and David’s Conversations) game formulated as a four-by-four asymmetrical SPS (Sir Philip Sidney) game: The terminal nodes show the payoffs (inclusive fitness) of Cindy and David, respectively. The dotted lines depict Cindy’s information sets, but David may not be able to distinguish the decision nodes connected by the dotted lines.

First, SPS game was invented by Maynard-Smith (1991) to model the reliability (honesty) of animal communication by narrating a well-known story about British poet-soldier Sir Philip Sidney (1554-1586), who was fatally injured in a battle, with the immortal words “*thy necessity is greater than mine*”, passed his water bottle to one fellow soldier who was also a casualty. Somewhat ironically, when transforming the story into the SPS game, Maynard-Smith (1991) introduced dishonest (cheating) possibilities in this heroic legend: Sir Philip’s comrade could cheat, *i.e*., requesting water bottle while he was not actually injured. On the other hand, Sir Philip could be selfless by donating the water bottle to his comrade or selfish by doing nothing. The selfish *vs*. selfless and dishonest *vs*. honest essentially introduce the cooperation and communication into SPS game, respectively. Since, to some extent, competition and cooperation can be considered as both sides of same coin, and communication can alter balance between competition and cooptation, or modulate the both, therefore, the SPS game spans the whole 3C processes (forces).

Second, SPS game was invented to demonstrate the handicap principle, which states that the signal of animal communication must be costly to be reliable (honest) and there is a strategic cost (the so-termed handicap) that ensures the honesty. For example, the peacock tail in flight is considered as a classic example of a handicapped signal of male quality, which signals the superior fitness of males to females in sexual selection. In human society, luxurious goods must be costly to be valuable (signaling social status). In the case of CDC game, there is a strategic cost (*c*) associated with miscommunication between Cindy and David. The SPS game not only provides a framework to model the strategic cost (*c*) capturing the possible noise communication between Cindy and David, but also offers measures to capture the costs associated with Cindy and David. For Cindy, there are two cost parameters (*a, b*) that are associated with underestimating or overestimating pandemic risk (*m*) level. For David, there is a cost parameter (*d*) that can capture the cost associated with David’s possible misjudgment. In one word, SPS game and more accurately extended SPS game model have four cost parameters (*a, b, c, d*) that can capture the costs associated with their potential faults and their miscommunications. For strategic modeling, this structure of cost parameters is sufficiently specific to capture major cost determinants but general enough for inferring equilibrium analytically.

Third, the extended SPS game has a relatedness parameter *k*, which is termed inclusive fitness (payoff) and measures the cooperation tendency between two game players. This fifth parameter (*k*) of CDC game is hence designed to measure cooperation (or competition) tendency in the policy-making system.

Fourth, as mentioned previously, there is a sixth parameter—the pandemic risk (*m*), which is the counterpart of injury probability of Sir Philip’s comrade in the classic SPS game. With the above-explained six parameters, the proposed CDC game is able to capture the estimation of pandemic *risk* (*m*), the *costs* (*a, b, c, d*) associated with reliability of the estimation, possible faults by Cindy and David, *benefits* (payoff) of the game strategies (analytically derived from the CDC game model). Finally, we have the relatedness parameter (*k*) that captures the intricacies of the strategic interactions between CDC chief and president. Although president may ignore chief’s advice for political consideration, they do have shared interests measured by *k*. With the six parameters, a system of 8 differential equations is formulated to describe the dynamics of the CDC game, and possible equilibriums of the dynamic systems for 16 strategy interactions can be analytically obtained and simulations can be performed. Using differential equations to formulate games as dynamic system rather than traditional matrix model is a huge advantage of evolutionary game, and allows for obtaining evolutionary stable strategies, which means that “mutation” of strategies with time are considered, and the strategies should be sustainable over time in general from a practical perspective.

Fifth, the SPS has an advantage over familiar PD game, namely it can simultaneously capture all three fundamental processes (forces) driving biological and/or social evolution, while the PD game can only capture two (competition and cooperation) excluding communication. For example, in the PD game, there are no direct communications between the two players (criminals) and they are questioned by police officer separately. In the SPS game, both players have explicit communications, and in fact, the game is a request-response game. Furthermore, the communicated information can be asymmetrical (deception). This difference makes SPS game a more appropriate choice than PD game for our research purpose.

Intuitively, and also in fact, there are arguable only four major kinds of anti-pandemic, public-health *policies* regarding lockdown or reopen, including (*i*) low-risk open & high-risk closed (LOHC), (*ii*) high-risk open & low-risk closed (HOLC), (*iii*) always open (AO), and (*iv*) and always closed (AC). These are four fundamental *policies* or practices a country (region) may adopt, and in this article we distinguish *policies* (referring to the above four) from *strategies*. Note that, although we use the term policy in this article since we believe the usage is more natural in the context of public-health policy-making, the four policies described above and referred in later sections, strictly speaking, should be termed as strategy or meta-strategy in the terminology of game theory.

With the CDC game, we postulate that the four policies should be mapped to the output of CDC game system, specifically, the ultimate decisions *made* by David with advice from Cindy. In other words, policies are the product or output of the decision-making process (system) such as the strategic CDC game. By distinguishing between the policy and strategy, one important gain is to shed light on the mechanism or process of decision-making process—how a particular policy can be reached by the advisor-decision-maker body such as CDC-President taskforce. Furthermore, one can also investigate the influences of various pandemic and/or socio-economic factors (as captured by the previous mentioned 6 parameters) on the policy-making process and outcome.

We admit that the *four-policy* classification is the result of somewhat simplified view, which is necessary as explained previously; nonetheless we argue that it is sufficiently general to cover the full spectrum of currently adopted anti-COVID policies in the world. At present, the LOHC policy is adopted by most countries (regions) in the world, while no country has adopted its opposite (*i.e*., HOLC), which is irrational if not anti-intelligent. A handful of countries have adopted *laissez-faire* strategy (passive herd immunity), which is equivalent to AO policy. The strict AC policy appears impractical and no country (region) has claimed to adopt such a policy. However, arguably, the dynamic zero-out (infections) adopted by a handful of countries such as China and New Zealand may be considered as either AC (in terms of effects) or LOHC (in terms of implementation).

## Methods: The CDC Game Model

### The conceptual formulation of CDC game model

After near two years of struggling against the COVID-19 pandemic, the vaccinations are bringing up, but the delta and emerging lambda are damping down, the hopes for reopening. Cindy, the CDC (center for disease control) chief scientist, is preparing her weekly briefing for the president, Mr. David Pearce, a newly elected president who has campaigned to bring peace and super humanity to the country and world. Cindy’s team has been evaluating the nation’s status of pandemic and vaccinations, and she realized that her team can only obtain a simple probability metric (*m*) to deliver her evaluation of the pandemic status, with *m* being worsen (high risk) and (1-*m*) alleviated (low risk). David, on the other hand, may need to balance the health *vs*. wealth, the public interests *vs*. other potentially conflicting interests. He may or may not follow Cindy’s advice with potential loss (*d*) depending on the reliability of Cindy’s advice and other factors on his side such as potential influences on economy and political correctness of his decision.

Furthermore, the trust between Cindy and David has been raising red light, and their cooperative relationship is far from perfect. For example, given that Cindy was appointed by previous administration and she might also be concerned with the risk of becoming a scapegoat in the case of catastrophic failure in anti-pandemic policy. In addition, Cindy’s aptitude may be imperfect and can be faulty. In extreme cases, Cindy may delay or even hide her evaluation of the true state-of-the-pandemic. Cindy’s potential faults, errors (benign faults) or dishonesty (malicious faults) can carry potential loss (cost) in the form of parameter *a* (underestimating risk *m*) or *b* (overestimate risk *m*), which influences her fitness or payoff.

Despite the potential communication issues between Cindy and David, both of them are certainly patriots and share common interests, and they tend to collaborate closely to maximize public and national interests. However, Cindy’s care of her reputation as a scientist and David’s care of his public approval rates are not necessarily always consistent, and may create competition. The balance between cooperation and competition between their interests can be measured by a *relatedness* parameter *k*, which is proportional to their tendency of cooperation. Furthermore, there is the cost (*c*) associated with strategic mistakes, the medical and socioeconomic consequences of misjudgment or miscommunication of the true state of pandemic, including consequences of possible cascading faults from both Cindy and David, *i.e*., cost of systemic failure.

Table 1 and Fig 1 exposed the formulation of the CDC (Cindy and David’s Conversations) problem as an extended SPS (Sir Philip Sydney) game, and Box S1 further explained the CDC game parameters. The so-termed extended SPS game refers to the transformation of the classic SPS game (Maynard-Smith 1991) from 2×2 to 4×4 asymmetrical game performed by Bergstrom & Lachmann (1997, 1998), Huttegger & Zollman (2010), Whitmeyer (2020, *etc*. In Table 1, the top section is the Sir Philip Sidney (SPS) game proposed by Maynard-Smith (1991) and further extended by Huttegger & Zollman (2010), and Whitmeyer (2020), as well as Ma & Zhang’s (2021) ABD game. In formulating the CDC game (the bottom section), we adopted slightly different symbols with the classic SPS game, with Cindy for advisor (signaler or message sender) and David for decision-maker (responder or message receiver). For the CDC game, table 1 listed the 4 possible options (strategies) Cindy and David each may have and their 16 (4×4) possible strategy interactions, mirrored 16 options of the CDC game or the 16 elements (cells) of the strategy matrix. Fig 1 further illustrates the CDC as a decision-tree and strategy matrix. The differential equation system for the CDC game is introduced in the next sub-section.

As exposed in Table 1, Fig 1, and Box S1, with the strategy *C*_*1*_, we map “open” (low risk) in the CDC to signaling (healthy) in the classic SPS, *i.e*., C1=“Cindy advises open only if she judges low-risk.” This is equivalent to map “closed” to “high risk.” Regarding David’s response, we map “closed” (high risk) in the CDC to “no signaling” in the original SPS, *i.e*., *D*_*1*_=“Closed only if Cindy advises closed.” Other strategy interactions can be mapped similarly.

As further exposed in Table 1 and Box S1, there are three elements in trans-formulating the classic SPS into CDC game, *who* (are the game players), *what* (messages are sent and responded), and *how* (the message is transmitted). From the classic SPS to the extended SPS games including our CDC game, the strategy set is expanded to 16 (4×4) strategy interactions from 4 (2×2) interactions. Besides the three elements, a few additional points are particularly worthy of reiterations (see Box S1 for the details). First, once the first cell (*C*_*1*_, *D*_*1*_) in the strategy matrix (set) is specified, the remaining 15 cells are set to keep consistent mathematic logic. Actually, once the “*who*” and “*what*” are set, the third element (“*how*”) may only influence the order of strategies in the strategic matrix, but not their mathematical logic. Second, although the classic SPS game is 2×2 asymmetric and not reciprocal, the extended SPS game including the CDC game is, in effects, equivalent to a *reciprocal* game given that virtually all 4×4 (16) possible strategy options are included in the strategy set (completeness). Due to this completeness or reciprocal equivalence, approximately ½ of the strategies may be unnatural or senseless (without discernible meaning or purpose), if the other ½ is considered as natural or rational. We take the advantages of those unnatural strategy interactions to capture the idiosyncrasies in decision-making for anti-pandemic, some of which could be not only counter-intuitive, but also unscientific, mirroring the various faults/noises in CDC explained previously.

In Table 1 and Fig 1, a series of labels (A-P) for the 16 (4×4) combinatorial strategies of Cindy and David are assigned to facilitate the discussion on the CDC game. Furthermore, the bottom section of Table 1 also list the four basic anti-pandemic policies introduced in the introduction section, and their corresponding equilibriums of the CDC game, but further explanation of the relationship between policies and equilibriums are delayed to the section of results.

### The differential equations of CDC game model

We adopt the extended SPS game model formulated by Huttegger & Zollman (2010) to describe the CDC game. The two-population replicator dynamics for Cindy and David, based on Huttegger & Zollman (2001), can be written as follows:

Let *x*_*i*_ be the relative frequency of Cindy’s strategy type *i*, and *y*_*j*_ be the relative frequency of David’s strategy type *j*, where *i, j*=1, 2, 3, 4, the replicator dynamics is written as follows:

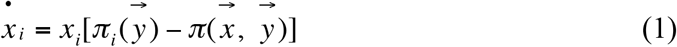

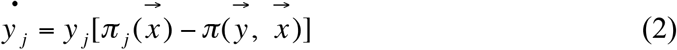

where 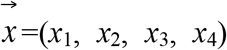 and 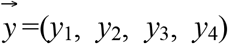, are the strategy sets of Cindy and David respectively; 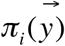 is the payoff of *i* strategy against 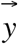 and 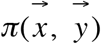 is the average payoff in Cindy’s population; 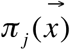 is the payoff of *j* strategy against 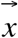 and 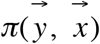 is the average payoff in David’s population.

## Results: Analysis and Simulation of the CDC game

### Equilibriums and Stabilities of the CDC Game

In this section, we take advantage of Huttegger & Zollman (2010) extensions and analytical results on the extended SPS game, and summarize the equilibriums of the CDC game in Table 2, and the payoff of the CDC game strategies in Table 3, respectively. From equations (1) & (2), six types of equilibriums are derived and summarized in Table 2, and a brief description is presented below:

**Table 2.** From CDC (Cindy and David’s conversations) game equilibriums/stability to anti-pandemic policies

| **Equilibrium Type**<br>(see Box S2 for definitions) | **Necessary conditions for the existence of the equilibriums**<br>(see Box S1 for CBD parameters) | **Stability Property**<br>(see Box S2 for definitions) | **Anti-pandemic Policy** (also see Table 1) |
| --- | --- | --- | --- |
| *A*—Nash equilibrium<br>(Apparently rational equilibrium) | $a > kd - c > b \ \& \ a > (d/k) > b$ | Asymptotically stable | LOHC (low-risk open and high-risk closed) |
| *F*—Signaling ESS (Separating equilibrium) | $a > c + kd > b \ \& \ a > (d/k) > b$ | ESS (Evolutionary Stable Strategy) and is Asymptotically stable. | LOHC |
| * *J* and *K* are behaviorally equivalent.<br>$(C_3, (1-\lambda)D_2 + \lambda D_3)$<br>—Pooling equilibrium | $d > k[ma + (1-m)b]$<br>& $\lambda \geq 1 - [c/(a - kd)]$ | Neutrally stable | AO (always open) |
| * *I* and *L* are behaviorally equivalent.<br>$(C_3, (1-\mu)D_1 + \mu D_4)$<br>—Pooling equilibrium | $d < k[ma + (1-m)b]$<br>& $\mu \geq [1 - c/(kd - b)]$ | Neutrally stable | AC (always closed) |
| *L*—Pooling equilibrium | $d < k[ma + (1-m)b]$ | Neutrally stable | AC |
| $(\lambda C_2 + (1-\lambda)C_4, \mu D_2 + (1-\mu)D_3)$<br>—Polymorphic Hybrid equilibrium<br>$\lambda = \frac{k[ma + (1-m)b] - d}{(1-m)(kb - d)} \ \&$<br>$\mu = \frac{c}{b - kd}$ | $a > (d/k) > b \ \& \ b - kd > c \ \&$<br>$d > k[ma + (1-m)b]$ | Lyapunov stable<br>(Asymptotically stable or unstable) | LOHC, AO, or AC |

\**J* & *K* mean that $(C_3, D_2)$ and $(C_3, D_3)$ are behaviorally equivalent. Similarly, *I* & *L* mean that $(C_3, D_1)$ and $(C_3, D_4)$ are behaviorally equivalent.

**Table 3.**
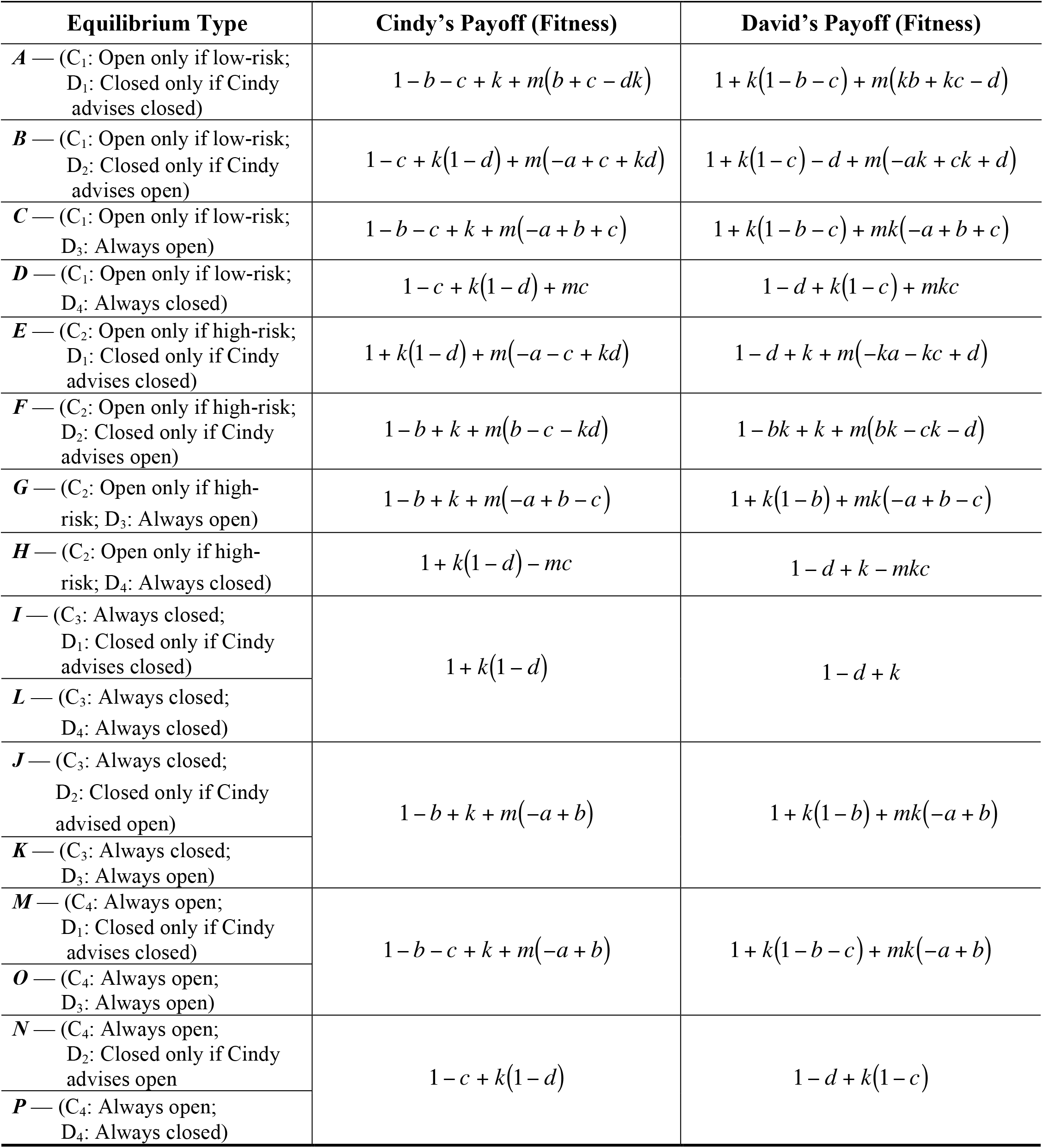
The expected payoffs for Cindy and David at the equilibriums of the CDC game

#### Strategy A has a Nash equilibrium and can be mapped to LOHC (low-risk open and high-risk closed) policy

With the strategy (pair or interaction) ***A*** of (*C*_1_ *vs. D*_1_), Cindy advises for *open* only if she perceives low risk of pandemic, and David decides *closed* only if Cindy advises for *closed*. In this strategic interaction, David follows Cindy’s advice, while Cindy seems rational with her strategy of “open only if low risk.” The strategy pair is Nash equilibrium and is asymptotically stable (*see* Box 2 for the terminology interpretations). Overall, the strategy pair ***A*** seems to be one of the most rational CDC strategy interactions from scientific perspective. Furthermore, since ***A*** is Nash equilibrium, nobody would regret for their decisions once they are committed. Strategy ***A*** appears to be the most natural and rational strategy.

Strategy ***A*** is nearly universally accepted by most countries in the world, and it is mapped to the number #1 anti-pandemic public health, *i.e*., the low (risk) open and high (risk) closed (LOHC) (Tables 1 and 2). The LOHC is arguably the most rational among the four fundamental policies. The LOHC policy can be made from strategy ***A*** with Nash equilibrium is obvious. However, the polymorphic hybrid strategy (PHS) can also lead to the LOHC policy, which may explain the idiosyncrasies occurred in various parts of the world regarding the anti-pandemic policy. That is, although the LOHC is adopted ultimately, the decision-making process to reach the policy could involve some irrational or unnatural strategy interactions, *e.g*., could be a trial-and-error process.

#### Strategy F is an ESS (evolutionary stable strategy), asymptotically stable, and can be mapped to LOHC policy

With the strategy pair *F* of (*C*_2_ *vs. D*_2_), Cindy appears irrational and advises for open if the risk is high, and perhaps she believes that the cost for closure is too high even though the pandemic risk is high. David appears more rational and overrules (reverses) the advice from Cindy, possibly either he does not trust Cindy’s advice or he believes that the benefits from closure outweigh the cost when the risk is high. Since ***F*** is an ESS, their decisions (behaviors) may be mutated when the pandemic expands and ultimately they may be converted from one strategy to another. Also because ***F*** is asymptotically stable, their consensus (equilibrium) has a region (range), which means both Cindy and David have certain level of flexibility in their behaviors. Overall, in this strategy interaction, the president (David) seems to have an intuitive and more rational attitude to the risk and decision than his advisor, Cindy.

Specifically, when the risk is high, Cindy would advise for open per *C*_2_ strategy, but David would choose to close per D_2_ strategy (David reverses Cindy’s recommendation). Hence, the strategy ***F*** again maps to the LOHC policy, as strategy ***A*** does. When the risk is low, Cindy offers no advice (her default option), but David has to make a decision anyway. Without advice from Cindy, he would choose his default option of open. That is, when no advice from Cindy, there should be an alternative to the scenario when Cindy advises him. With this logic, David’s decision would still lead to rational LOHC policy. Therefore, the strategy interaction ***F*** can evolve to an evolutionary stable strategy, and be mapped to the LOHC policy ultimately.

Therefore, both strategy ***A*** and ***F*** interactions can generate the LOHC policy: in the case of ***A*** strategy interaction, David usually simply follows Cindy’s advice, while in the case of ***F***, David usually overrule Cindy’s advice.

#### Strategy L is a pooling equilibrium and can be mapped to AC (always closed) policy

With the strategy pair ***L*** of (*C*_3_ *vs. D*_4_), Cindy always advises closure regardless of the risk level, and David always decides closure regardless of Cindy’s advice. The equilibrium is neutrally stable. From a pure theoretic perspective, the equilibrium type ***L*** is more likely to become established because it has a larger basin of attraction, which measures the likelihood of evolving under standard evolutionary dynamics [Eqn. (1)-(2)]. From a pure health perspective without considering wealth, the strategy can be optimum. Overall, the strategy pair appears to mirror a pair of advisor and decision-maker who take totally same attitudes to the disease and risk. Since the equilibrium of strategy is a pooling equilibrium, Cindy’s has no influences on David although Cindy’s advice is always consistent with David’s decision.

Obviously, this strategy interaction can be mapped to AC (always closed) policy for anti-pandemic. This strategy may be an optimum if the pandemic is over within a reasonably short period or economic loss is not a concern. Furthermore, if pandemic can be suppressed in a short period, the economic loss would not be a concern either.

#### Mixed Strategies of J and K are pooling strategies and both are behaviorally equivalent, and can be mapped to AO (always open) policy

When Cindy adopted strategy *C*_3_ (always closed), and David adopted a mixed strategy of *D*_2_ (Closed only if Cindy advises open) with probability (1−λ) and *D*_3_ (always open) with probability λ, there is a pooling equilibrium that is neutrally stable. The condition for the equilibrium to occur is *d* > *k*[*ma* + (1 − *m*)*b*]. David’s mixed strategy is (1 − *λ*)*D*_2_ + *λD*_3_, and *λ* ≥ [1 − *c* /(*a* − *kd*)] determines the strategy of David.

Theoretically, when David hesitates, he can resort to λ to make an optimal decision, *i.e*., choose *D*_*2*_ when *λ* ≥ [1 − *c* /(*a* − *kd*)], or *D*_*3*_ when *λ* < [1 − *c* /(*a* − *kd*)]. Since Cindy always advises for closure, David would always decide to open. Strategies ***J*** and ***K*** are behaviorally equivalent with each other.

A scenario for achieving this equilibrium can be like this: Cindy always advises for closure regardless of the risk, David may hesitate but in the end he still ignores Cindy’s advice and keep open. This scenario maps to AO (always open) policy. Herd immunity strategy could be considered as belonging to this strategy. In addition, the equilibrium of mixed ***J*** & ***K*** strategies is pooling, which implies that David’s decision is ‘dictatorship’ style. That is, although he may hesitate, but ultimately still ignores Cindy’s advice.

#### Mixed strategies of I and L are pooling strategies and both are behaviorally equivalent, and can be mapped to AC (always closed) policy

When Cindy adopted strategy C_3_ (always closed) and David adopted a mixed strategy of D_1_ (Closed only if Cindy advises so) with probability (*1*−*μ*) and D_4_ (always closed) with probability μ, where *μ* = 1 −[*c* /(*kd* − *b*)], there is a pooling equilibrium that is neutrally stable. Similar to previous *J* & *K* strategies, there is a pooling equilibrium that is neutrally stable, but here *μ* determines the strategy of David. Intuitively, since Cindy always advises closed, David may hesitate: but in the end, he still decides to close. The condition to determine David’s behavior seems to be opposite with the previous mixed strategy interaction of *J* & *K*. Strategies ***I*** and ***L*** are behaviorally equivalent with each other.

In practice, similar to strategy *L*, this mixed strategy of *I* and *L* maps to AC (always closed). The policy should be optimum when pandemic ends in a short period (“wealth” is not a concern) or economic loss from closure is not a concern.

#### *Polymorphic hybrid strategies* (PHS)

Cindy adopted a mixed strategy of *C*_2_ (open only if high-risk) and *C*_4_ (always open), and David also took a mixed strategy of *D*_2_ (closed only if Cindy advises open) and *D*_3_ (always open). With these options, there are hybrid equilibriums that are *polymorphic*. Overall, Both Cindy and David may hesitate, but they seem partially rational in the sense that Cindy may consider risk level and David may follow Cindy’s advice. For Cindy, she may recommend open or closed, and for David he may decide open or closed depending on the conditions explained below. This obviously is the most sophisticated type of equilibrium.

First, how would Cindy make her recommendation? In fact, it is the parameter μ that determines her recommendation to choose either *C*_2_ or *C*_4_. Theoretically, when she hesitates, she can resort to μ to make an optimal decision, *i.e*., choose *C*_2_ when *μ* > *c* /(*b* − *kd*), *C*_4_ when *μ* < *c* /(*b* − *kd*). When *μ* = *c* /(*b* − *kd*), she tends to adopt mixed strategy *λC*_2_ + (1 − *λ*)*C*_4_.

For example, when *μ* > *c* /(*b* − *kd*), Cindy would prefer to choose C_2_, she would advise open even if high risk. This choice would suggest that Cindy is skeptic of closure. In contrast, when *μ* < *c* /(*b* − *kd*), Cindy would advise open anyway.

Second, how would David make his decision of *D*_2_ or *D*_3_ in response to Cindy’s advice? David’s decision would depend on parameter λ. When *λ* > {*K*[*ma* + (1 − *m*)*b*] − *d*}/[(1 − *m*)(*kb* − *d*)], David would prefer to choose D_2,_ (closed only if Cindy advises open, *i.e*., overrule or negate Cindy’s advice), or D_3_ (always open) when *λ* < {*K*[*ma* + (1 − *m*)*b*] − *d*}/[(1 − *m*)(*kb* − *d*)]. When *λ* = {*K*[*ma* + (1 − *m*)*b*] − *d*}/[(1 − *m*)(*kb* − *d*)], he tends to adopt mixed strategy *μD*_2_ + (1 − *μ*)*D*_3_. This hybrid equilibrium is Lyapunov stable, which implies that the actual stability may vary depending on the values of parameters. In other words, the stability of equilibriums may be broken when conditions such as risk level and/or cost change dramatically. Fig 2 shows an example of the decision-making in the phase space of the polymorphic hybrid equilibrium, obtained from the simulation study explained below. The polymorphic hybrid strategies, arguably, represent the most sophisticated options, which reflect the dynamic (evolutionary) nature of the masking-or-not decision-making.

**Fig 2.**
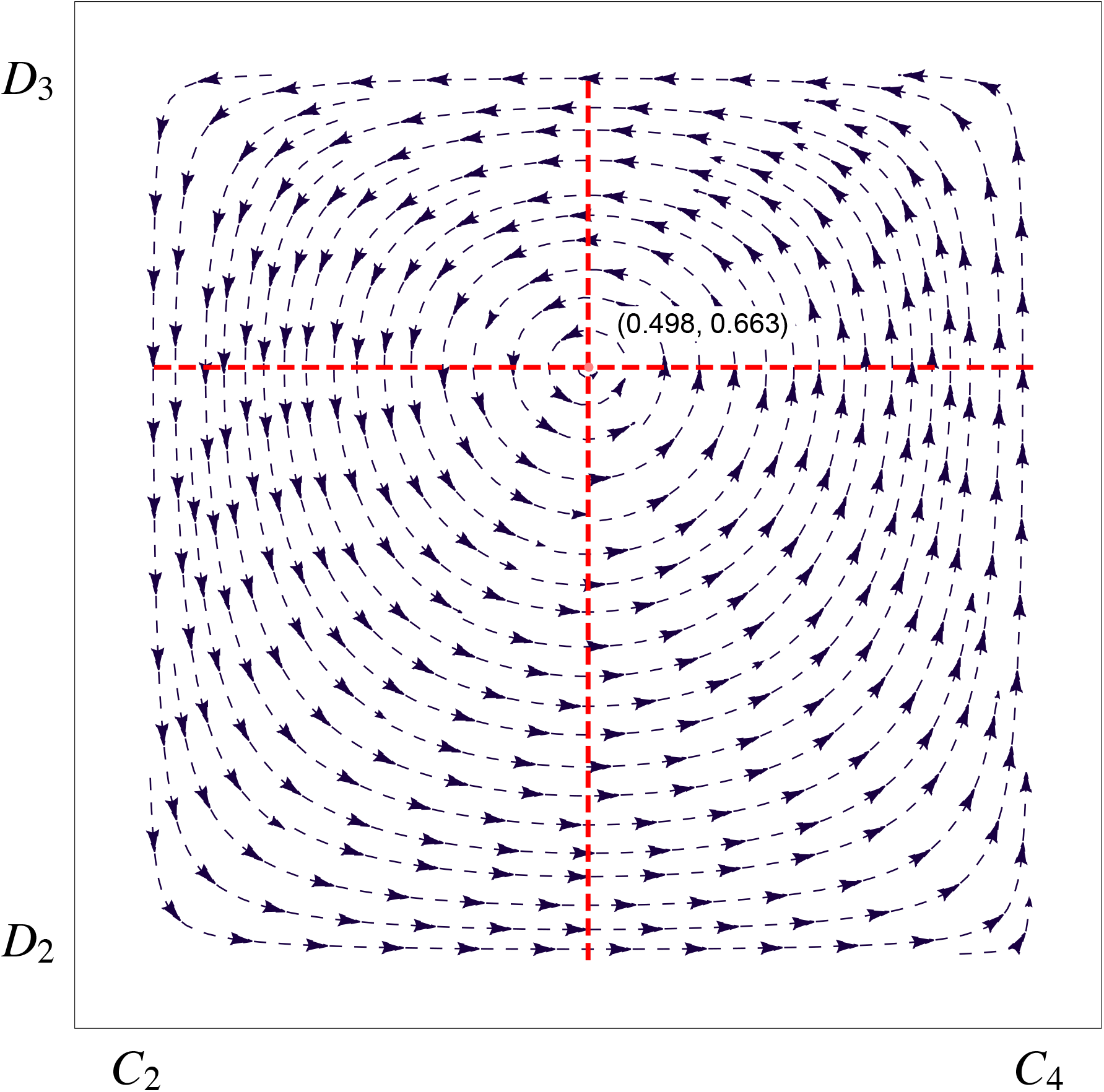
An example output from the simulation of CDC game, *i.e*., the phase portrait of the polymorphic hybrid equilibrium (PHE) with the hybrid strategy of Cindy (*C*_2_, *C*_4_) *vs*. David (*D*_2_, *D*_3_) under following parameter values: *a*=*0.7, b*=*0.3, c*=0.1, *d*=0.3, *m*=*0.6* and *k*=0.5. Cindy and David make their decisions based on (*A, μ*)=(0.498, 0.663). If *μ>0.663*, Cindy prefers to choose *C*_*2*_ over *C*_*4*_, as indicated by the direction of portrait arrow (to left). If *μ<0.663*, Cindy prefers to choose *C*_*4*_ over *C*_*2*_, as indicated by the direction of portrait (to right). When *μ=0.663*, the *mixed strategy of C*_*2*_ and *C*_*4*_ is chosen. Similarly, David makes his strategic decisions based on the value of parameter *A >0.498, <0.498 or =0.498*.

In practice, the *polymorphic hybrid strategy* (PHS) may lead to three of the four policies, except for irrational HOLC (high-risk open & low-risk closed). For example, with (*C*_2_, *C*_4_) and (*D*_2_, *D*_3_) strategy interactions, any of the four strategy interactions (*F, G, N, O*) can happen. If *F* is chosen, then it may lead to LOHC; if *G* or *O* is chosen, then it may lead to AO; if *N* is chosen, it may lead to AC. Note that none of the polymorphic strategies with equilibriums can be mapped to HOLC, which indicates that the HOLC is so irrational and is unlikely to sustain (reaching equilibrium) in practice.

In summary, as listed in Table 1 (the bottom section) and Table 2 (the last column), arguably the most rational LOHC policy can be the output of equilibriums ***A*** & ***F***; the AO policy can be the output of equilibriums ***J*** & ***K***; the AC policy can be the output of equilibriums ***I*** & ***L*** or ***L***. The polymorphic hybrid strategy (PHS) can produce any of the three previously discussed policies, *i.e*., LOHC, AO, or AL. However, none of the six types of equilibriums can produce the HOLC policy, which should be expected that given the policy is unlikely to sustain in practice.

#### Strategy interactions without equilibriums

For strategy interactions that do not produce equilibriums, there are the following mappings: strategy interactions *D,H,L,P,N*, and *I* can produce AC policy, and strategy interactions *C,G,K,O, M*, and *J* can produce AO policy. Since these strategy interactions are not evolutionarily stable, which implies that they are not sustainable in practice, we do not discuss them further in this study.

Finally, strategies ***B*** & ***E*** can be mapped to irrational HOLC policy, but both the strategies do not have any equilibrium (Table 2), which should be expected and is consistent with the reality that the HOLC policy is so irrational and is unlikely to sustain practically.

### Simulation Experiments

#### Simulation Procedures and Phase Portrait

Since the analytic solutions (Tables 2 and 3) are less intuitive in interpreting the decision-making process of CDC game, particularly, in understanding the influences of various CDC parameters (cost/benefit factors, risk level, cooperation tendency), we performed simulation experiments based on the replicator dynamics (Eqns. 1-2). Tables S1-S4 contain the 4 lists of equilibriums corresponding to the three public health policies (LOHC, AO, AC), and the list of PHS, which may be mapped to one of the previous three policies, depending on the specific parameter values of the CDC game. The simulations were performed based on the replicator dynamics (Eqns. 1-2) with a program developed by Ma & Zhang (2021). Specifically, the equilibriums were simulated and determined based on the necessary conditions (listed in Table 2) and corresponding payoffs were computed based on the equations listed in Table 3, with simulation step length=*0.1* for all of the model parameters (*a, b, c, d, k, m*), all of which range from *0* to *1*. A total of *10*^*8*^ simulations were performed to obtain the results, and the results were organized in terms of the policy category (online supplementary Tables S1-S4).

The strength of simulations helps us to illustrate the complex interactions between advisor and decision-maker intuitively, but the simulations cannot be exhaustive due to the continuous nature of the CDC model. In the following, we use the simulations to primarily address two questions: (*i*) The influences of various CDC parameters on the outcome (policies) of CDC game; those parameters approximate the cost/benefits, risk level, faults, miscommunications (dishonesty), *etc* in the decision-making system. (*ii*) Likelihood of each policy—the estimated probability of the policies based on *10*^*8*^ times of simulations. Before addressing the two questions, we demonstrate the simulation of PHS with a phase portrait (Fig 2) to illustrate possible interactions between Cindy and David in a hybrid strategy.

Fig 2 is plotted to demonstrate the dynamic properties of the PHS equilibrium, *i.e*., the phase portrait of the PHE (polymorphic hybrid equilibrium) with the hybrid strategy of Cindy (*C*_*2*_, *C*_*4*_) *vs*. David (*D*_*2*_, *D*_*3*_) under following parameter values: *a*=*0.7, b*=0.3, *c*=0.1, *d*=0.3, *k*=0.5, and *m*=*0.6*. David and Cindy reach their decisions based on (*λ, μ*)=(0.498, 0.663), which is an equilibrium point as displayed as red dot in Fig 2. When *μ>0.663*, Cindy prefers to choose *C*_*2*_ over *C*_*4*_, as indicated by the direction of portrait arrow (to left). When *μ<0.663*, Cindy prefers to choose *C*_*4*_ over *C*_*2*_, as indicated by the direction of portrait (to right). When *μ=0.663*, the *mixed strategy of C*_*2*_ and *C*_*4*_ is chosen. Similarly, David makes his strategic decisions based on the value of parameter *λ>0.498, <0.498 or =0.498*.

#### Influences of the cost, risk & cooperation/communication factors on the policies

##### Influences of Cindy’s cost parameter a on the occurrences of the policies

Fig 3A plotted the estimated probabilities of adopting three policies (LOHC, AO, AC) from *10*^*8*^ simulations of the CDC game under different cost parameter *a* (underestimating the risk) of Cindy. With the rise of Cindy’s cost (*a*) to underestimate the risk (*m*), the probability of adopting AO increases initially and reaches an asymptote under intermediate cost level (*a*≈0.3-0.5), and then decline steadily; that of adopting AC exhibited an opposite trend; and that of adopting LOHC policy is rather low but stable. The probability from adopting a policy generated by PHE (polymorphic hybrid equilibrium) is negligible. These probability trends appear natural. For example, when the cost (*a*) is relatively small (*a*<0.3), AO policy is favored (AC is disfavored), but when the cost is large (*a*>0.6), the trend is reversed. The cost parameter seems to have little influence on the likelihood of LOHC policy except when the cost is extremely high (*a*>0.7 approximately). The explanation for the negligible PHE probability is likely due to unduly complexity in implementing the PHE strategy. Indeed, we postulate that the small-probability associated with LOHC and PHE is due to their unduly complexities.

**Fig 3.** The probabilities of adopting three basic policies (LOHC, AO, AC) estimated from 10^8^ simulations of CDC game dynamics under different CDC parameter values; the fourth item “Hybrid” (*i.e*., PHE=polymorphic hybrid equlibrium) may be classified into one of the three previously listed basic policies, depending on complex interactions between Cindy and David; however, its probability and practical implications are negligible. *See* explanations below for specific **Figs 3A-3F**.

**Fig 3A.**
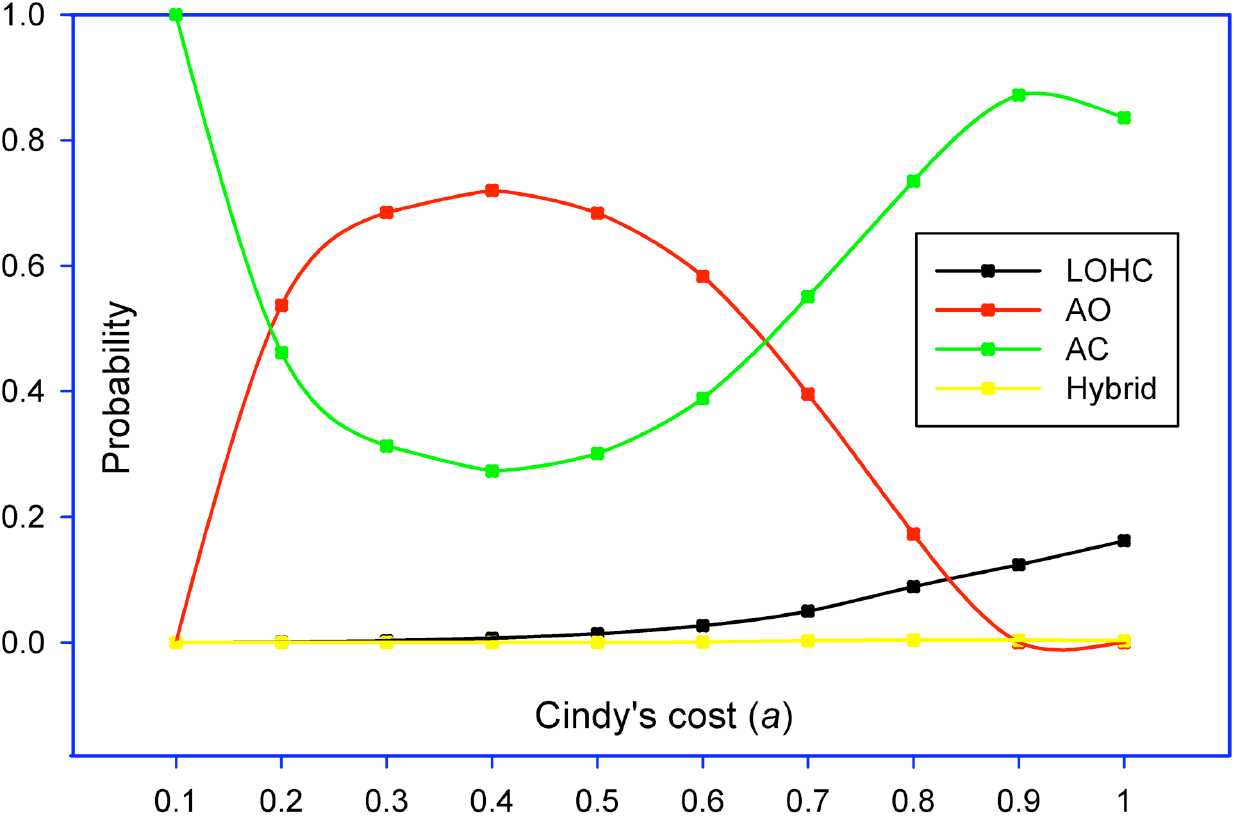
The probabilities of adopting three policies (LOHC, AO, AC) estimated from 10^8^ simulations of CDC game under different *cost* parameter (*a*) of Cindy. With the rise of Cindy’s cost (*a*) to underestimate the risk, the probability of adopting AO increases initially and reaches an asymptote at intermediate-risk level, and then declines steadily; that of adopting AC exhibits an opposite trend; and that of adopting LOHC policy is rather low but stable. The probability from adopting a policy generated by the hybrid strategy (*i.e*., PHE) is negligible.

**Fig 3B.**
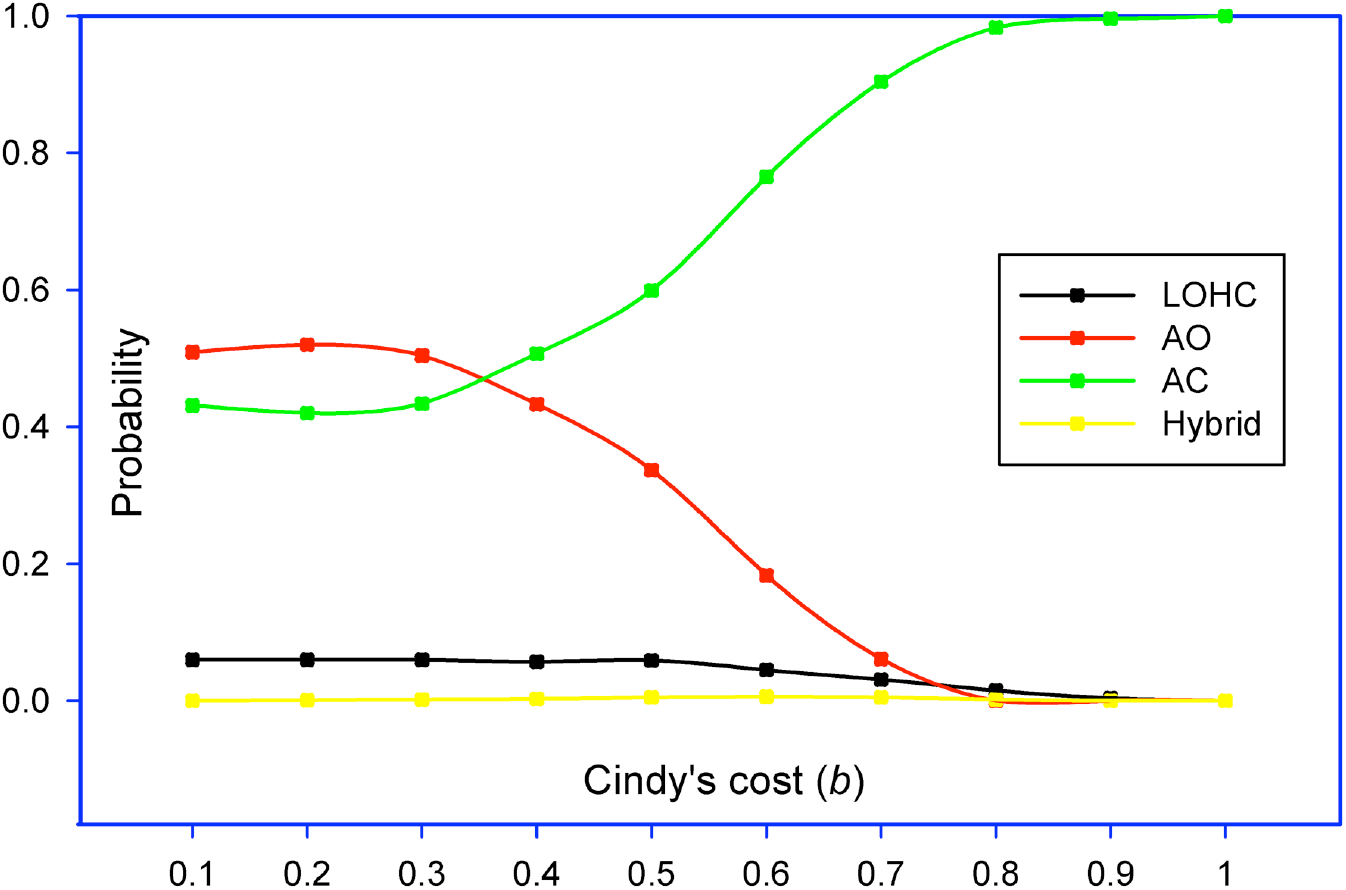
The probabilities of adopting three policies (LOHC, AO, AC) estimated from 10^8^ simulations of CDC game under different *cost* parameter (*b*) of Cindy. With the rise of Cindy’s cost (*b*) to overestimate the risk, the probability of adopting AC increases gradually and reaches an asymptote; that of adopting AO exhibits an opposite trend; and that of adopting LOHC policy is rather low but stable. The probability from adopting a policy generated by the hybrid strategy (*i.e*., PHE) is negligible.

**Fig 3C.**
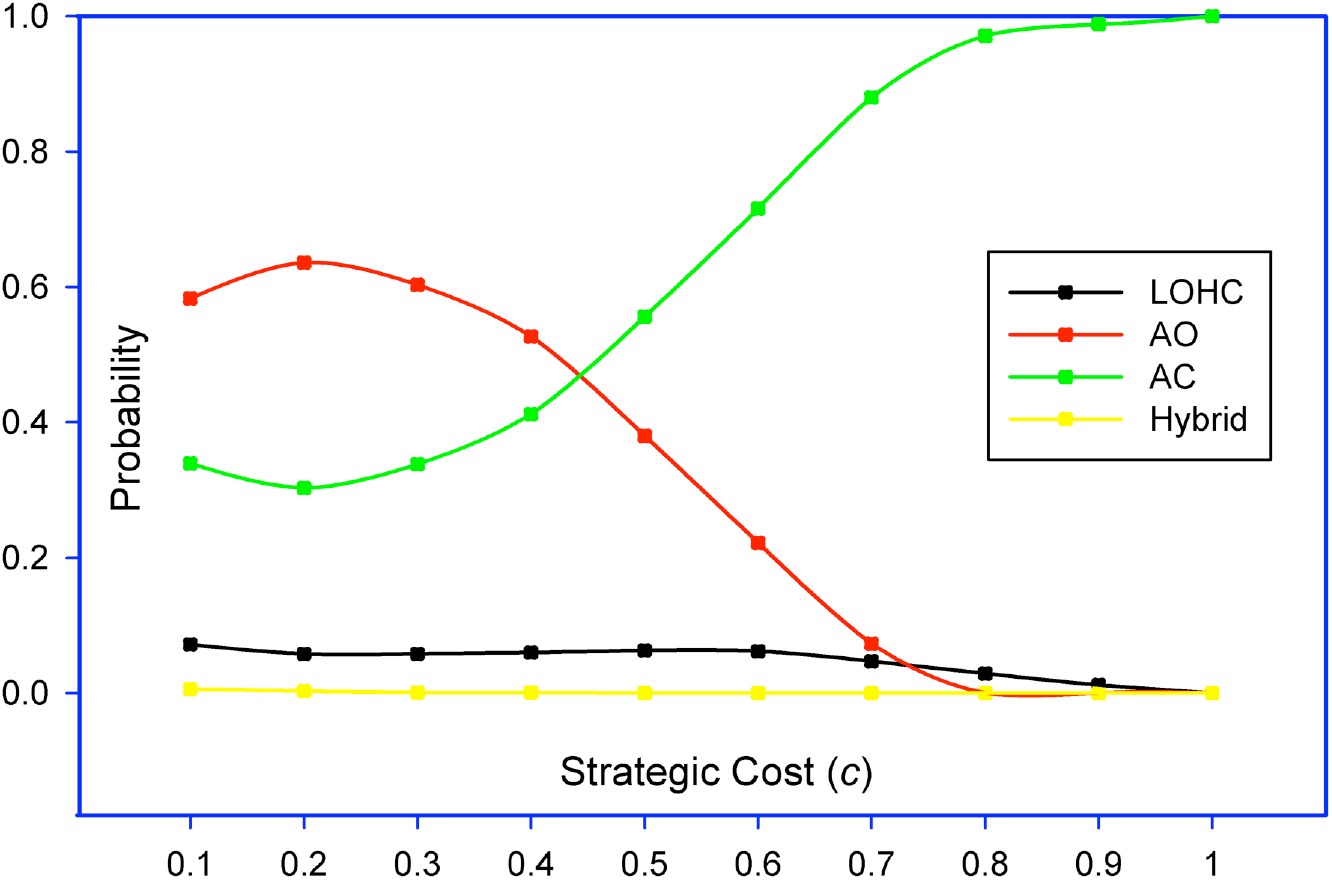
The probabilities of adopting three policies (LOHC, AO, AC) estimated from 10^8^ simulations of CDC game under different values of *strategic cost* (*c*): With the rise of strategic cost (*c*), the probability of adopting AC increases steadily and reaches an asymptote; that of adopting AO exhibits an opposite trend; and that of adopting LOHC policy is rather low but stable. The probability from adopting a policy generated by the hybrid (*i.e*., PHE) strategy is negligible.

**Fig 3D.**
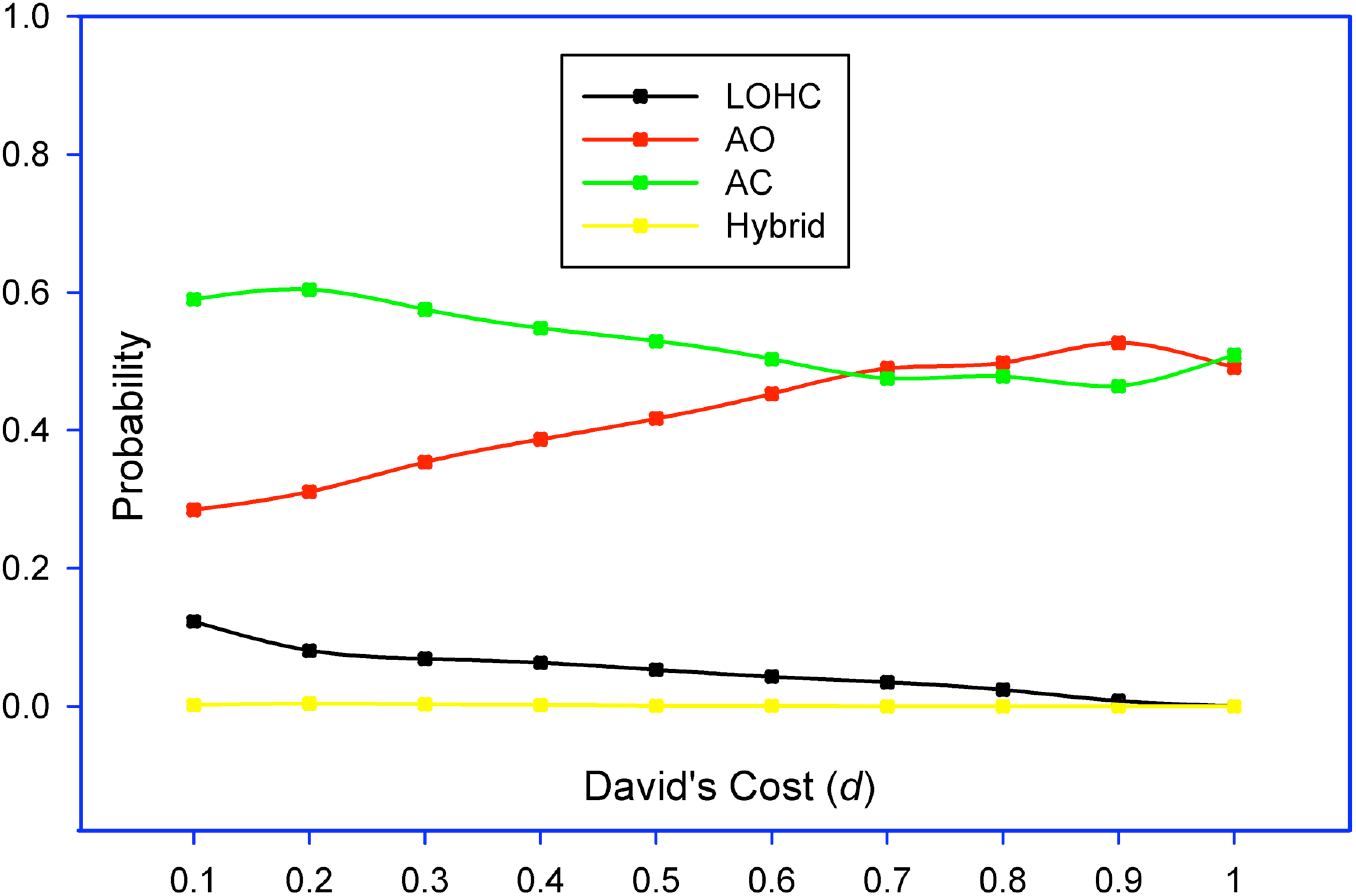
The probabilities of adopting three policies (LOHC, AO, AC) estimated from 10^8^ simulations of CDC game under different values of David’s *cost* (*d*): With the rise of David’s cost (*d*), the probability of adopting AO increases slowly and but declines when the cost is exceptionally high (>0.9); that of adopting AC exhibits an opposite trend; and that of adopting LOHC policy is low and decline slightly. The probability from adopting a policy generated by the hybrid strategy (*i.e*., PHE) is negligible.

**Fig 3E.**
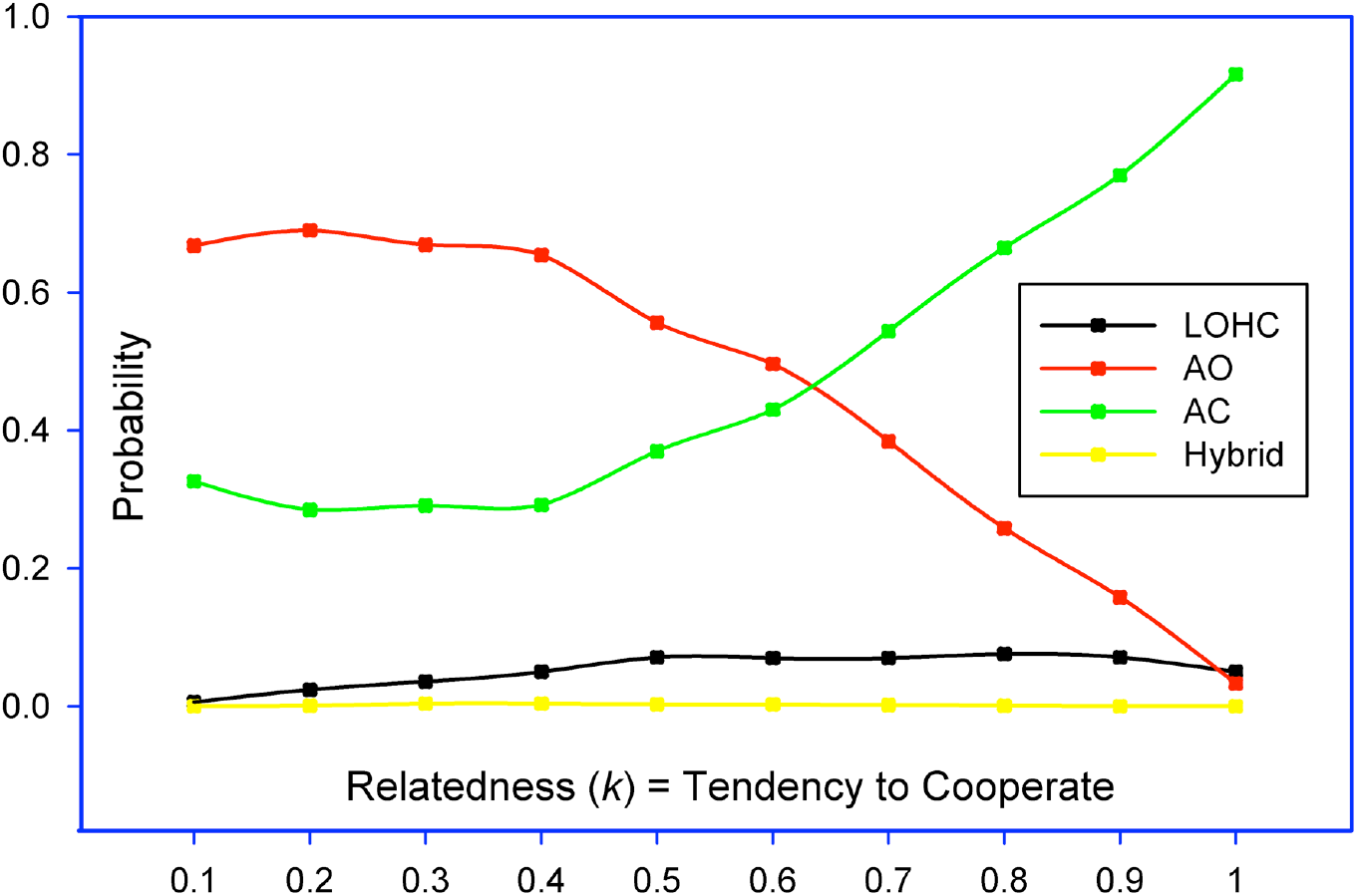
The probabilities of adopting three policies (LOHC, AO, AC) estimated from 10^8^ simulations of CDC game under different *relatedness* parameter (*k*): With the increase of *relatedness* (*k*) between Cindy and David (their collaboration tendency), the probability of adopting AO declines steadily; that of adopting AC increases gradually; and that of adopting LOHC is low but largely stable. The probability from adopting a policy generated by the hybrid strategy (*i.e*., PHE) is negligible.

**Fig 3F.**
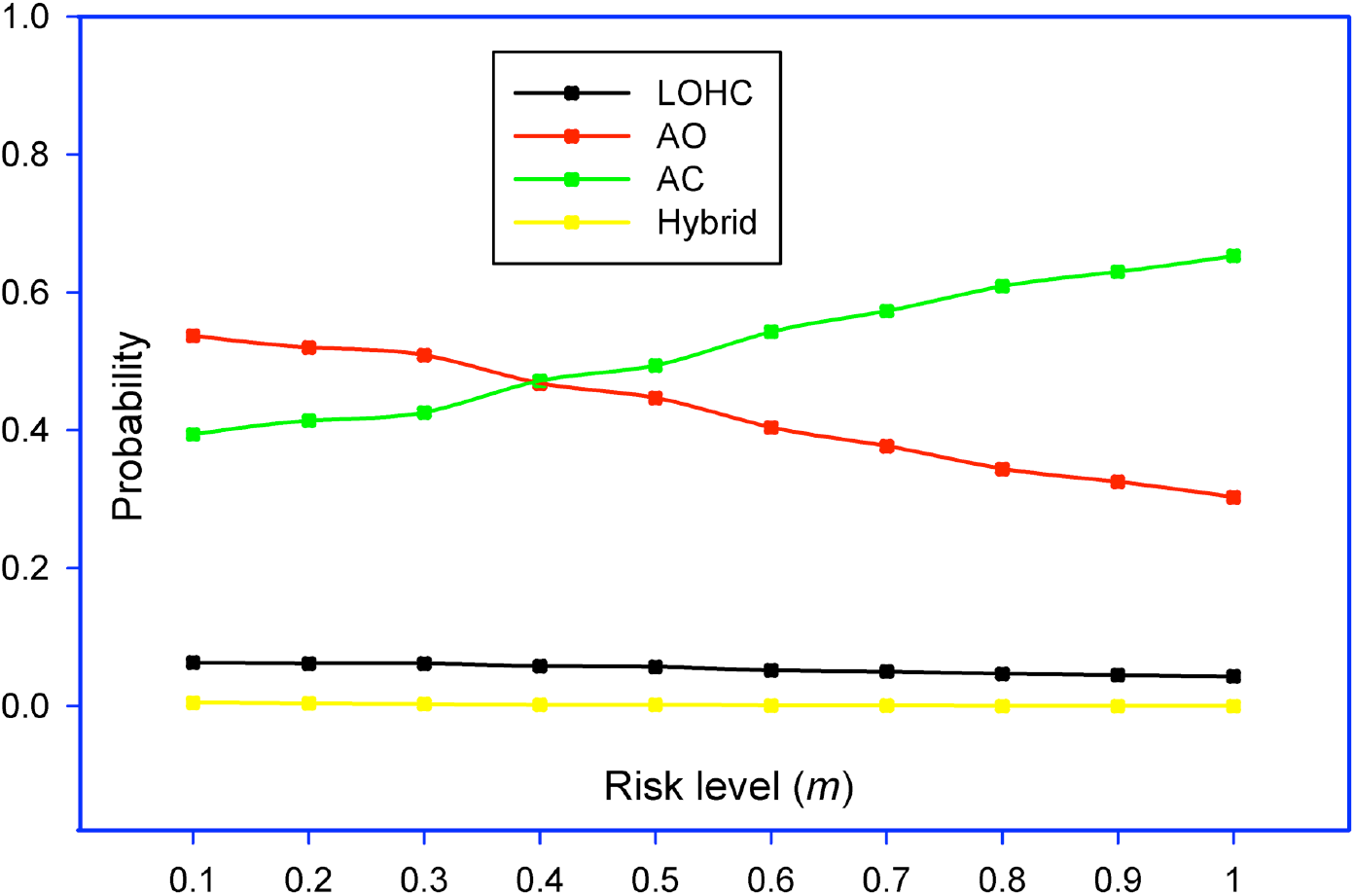
The probabilities of adopting three policies (LOHC, AO, AC) from 10^8^ simulations of CDC game under different risk level: With the rise of risk level (*m*), the probability of adopting AO declines steadily; that of adopting AC increases gradually; and that of adopting LOHC is low but stable. The probability from adopting a policy generated by the hybrid strategy (*i.e*., PHE) is negligible.

##### Influences of Cindy’s cost parameter b on the occurrences of the policies

Fig 3B plotted the estimated probabilities of adopting three policies (LOHC, AO, AC) from 10^8^ simulations of the CDC game under different cost parameter *b* (overestimating the risk) of Cindy. With the rise of Cindy’s cost to overestimate the risk (*b*), the probability of adopting AC increases gradually and reaches an asymptote; that of adopting AO exhibited an opposite trend; and that of adopting LOHC policy is rather low but stable. The probability from adopting a policy generated by PHE is negligible. These probability trends appear natural. For example, when the cost (*b*) of overestimation increases, AC policy is increasingly favored, and AO policy is increasingly disfavored.

##### Influences of strategic cost (c) on the occurrences of the policies

Fig 3C plotted the influence of strategic cost (*c*) on the likelihoods of the three policies (LOHC, AO, AC). With the rise of strategic cost (*c*), the probability of adopting AC increases steadily and reaches an asymptote; that of adopting AO exhibited the opposite trend; and that of adopting LOHC policy is rather low but stable. When the cost (*c*)>0.6, the likelihood of adopting LOHC declines slightly, which may have to do with the possibly rising difficulty in effectively implementing the LOHC policy. The probability from adopting a policy (which could be any of the three policies, AC, AO, LOHC) generated by PHE (polymorphic hybrid equilibrium) is negligible, and little influenced by the strategic cost (*c*). These probability patterns again seem natural. For example, with the rise of strategic cost (*c*), the policy becomes more conservative and increasingly in favor for AC and against AO.

##### Influences of David’s cost (d) on the occurrences of the policies

Fig 3D illustrated the influence of David’s cost parameter (*d*) on the probabilities of three policies (LOHC, AO, AC) from 10^8^ simulations of the CDC game. With the rise of David’s cost (*d*), the probability of adopting AO increases slowly, but declines when the cost is exceptionally high (>0.9); that of adopting AC exhibited the opposite trend; and that of adopting LOHC policy is low and decline slightly. The probability from adopting a policy generated by PHE (polymorphic hybrid equilibrium) is negligible. When David’s cost is exceptionally high (*d*>0.9), the policy is likely to reverse, which seems rather natural.

##### Influences of relatedness (k) on the occurrences of the policies

Fig 3E illustrated the influence of relatedness parameter (*k*) on the probabilities of adopting three policies (LOHC, AO, AC) from 10^8^ simulations of the CDC game. With the increase of relatedness (*k*) between Cindy and David (their collaboration tendency), the probability of adopting AO declines steadily; that of adopting AC increases gradually; and that of adopting LOHC is low but largely stable. The probability from adopting a policy generated by PHE (polymorphic hybrid equilibrium) is negligible. The finding seems natural again. For example, when the collaboration tendency increases, the AC policy is favored increasingly.

##### Influences of risk level (m) on the occurrences of the policies

Fig 3F illustrated the influence of pandemics risk (*m*) on the probabilities of adopting three policies (LOHC, AO, AC) from 10^8^ simulations of the CDC game. With the rise of risk level (*m*), the probability of adopting AO declines steadily; that of adopting AC increases gradually; and that of adopting LOHC is low and almost constant. The probability from adopting a policy generated by PHE (polymorphic hybrid equilibrium) is negligible. When the risk level *m*=0.4, the probabilities of AC and AO crossover with each other, at which both policies are equally likely.

#### Overall likelihoods (probabilities) of the three policies (LOHC, AO, AC)

In the previous section, we discussed the influences of single factor (CDC parameter) on the policy-making. In this section, our focus is on the overall probabilities of the three policies. In Fig 4, we plotted the overall probabilities of adopting three basic policies (LOHC, AO, AC), respectively, across all simulated parameter settings (*i.e*., averaged from the values across all parameters or Fig 3A-3F). The fourth item—PHE—can be classified into one of the three basic policies, depending on complex interactions between Cindy and David. However, to classify each PHE into one of the three policies, additional ‘real-time’ (*i.e*., the threshold value of parameter *λ* or *μ* determines the choice of mixed strategy) interaction information is necessary, which requires additional extensive simulations. Since the total (combined) probability (*P*=0.002) of the three policies from PHE is negligible, the lack of further classifications should only have negligible biases on the probabilities of the three policies drawn in Fig 4.

**Fig 4.**
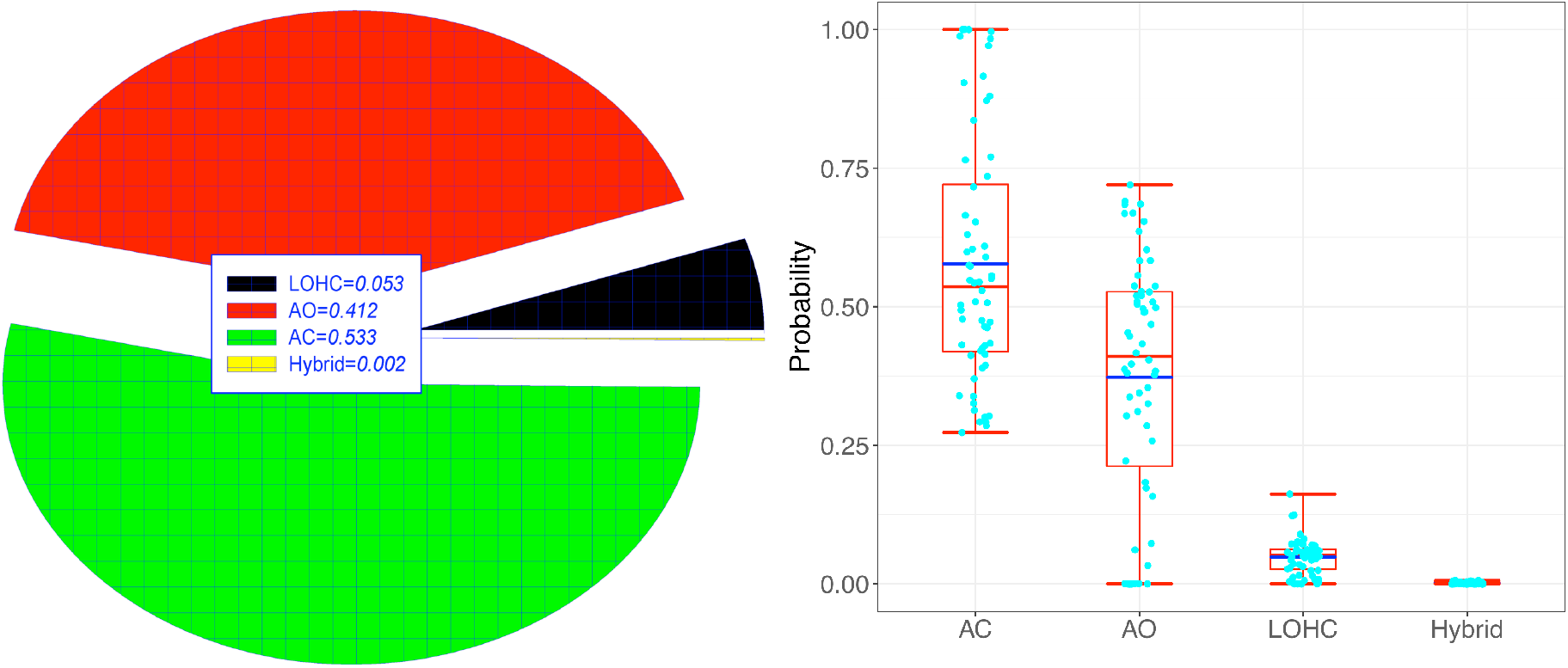
The probabilities of adopting three basic policies (LOHC, AO, AC) estimated from 10^8^ times of simulations of the CDC game *across* all possible parameter settings (*i.e*., averaged from the values across Fig 3A-3F). The fourth item—hybrid (*i.e*., PHE)—can be classified into one of the three basic policies, depending on complex interactions between Cindy and David; nevertheless, its probability is negligible and so is its practical implications. Fig 4A (pie chart) shows the ‘partition’ of the probability space (‘pie’). Fig 4B shows standard box chart, with blue-colored line representing for the mean, red-colored line for the median, top box red-line for 75% points, bottom box red-line for 25% points, as well as the minimum (bottom red-line) and maximum (top red-line). The insights from the CDC game simulation are clear: (*i*) The universally accepted LOHC (low-risk open and high-risk closed) appears ideal but hardly implementable in practice, as evidenced by slightly larger than the probability of small event (*P*=0.053). (*ii*) Highly complex polymorphic hybrid strategy (PHS) interactions seem promising, which theoretically can generate any of the three strategies (LOHC, AO, AC) depending on complex parameter setting (*i.e*., strategy interactions between Cindy and David), but the estimated probability is negligible (*P*=0.002). (*iii*) The extensive simulations from the CDC game suggest that KISS (keep it simple strategically and persistently) should be large-probability events, as indicated by the rather high probabilities of AO (always open: *P*=0.412) and AC (always closed: *P*=0.533) policies. (*iv*) Finally, the HOLC (High-risk open & low-risk closed) policy, which is not displayed here, is obviously anti-science and unnatural, and it may be generated from strategies B & E (Table 1), but they do not have equilibriums, which means unstable evolutionarily and is unlikely to sustain in practice.

From Fig 4, findings on the overall likelihoods (probabilities) of the three policies include: (*i*) The near universally adopted LOHC, *i.e*., a middle-ground of AC and AO policies, intuitively should be preferred, but can be too complex to enforce practically, as suggested by the only slightly larger than the probability of small event (*P*=0.053). (*ii*) Highly complex polymorphic hybrid strategy (PHS) interactions seem promising, which theoretically can generate any of the three strategies (LOHC, AO, AC) depending on complex parameter setting (strategy interactions between Cindy and David), but the estimated probability is negligible (*P*=0.002). (*iii*) The extensive simulations (*10*^*8*^ times) of the CDC game suggest that KISS (keep it simple strategically) should be large-probability events, as indicated by the rather high probabilities of AO (always open: *P*=0.412) and AC (always closed: *P*=0.533) policies. (*iv*) Finally, the HOLC (high-risk open & low-risk closed) policy, which is not displayed in Fig 4, is obviously anti-science and unnatural. Although LOHC may be generated from strategies B & E (Table 1), both strategies B & E do not have equilibriums, which means that they are evolutionarily unstable and unlikely to sustain in practice.

#### Payoffs of the CDC game players

In previous sections, our discussions are focused on the simulation of the influences of various CDC game parameters (that represent for various costs/benefits, risk and cooperation/ communication factors) on the strategies (policies). In this section, our focus is the payoffs of CDC game. Fig 5 plotted the payoffs of Cindy and David under different policies, respectively. Notably, the order of payoffs is different for Cindy (Fig 5A) and David (Fig 5B). With Cindy’s payoff, the descending order (from left to right) is AC>LOHC>AO>Hybrid, and with David’s payoff, the descending order is LOHC>AO>AC>Hybrid. The orders are made based on statistically significant differences based on Wilcoxon tests (*P*-value<0.05) (See Table S2). The opposite payoff-orders suggest that Cindy prefers tighter AC policy, while David prefers more relax AO policy. This difference may be due to the reality that David should be more concerned with the health *vs*. wealth tradeoff or dilemma.

**Fig 5.**
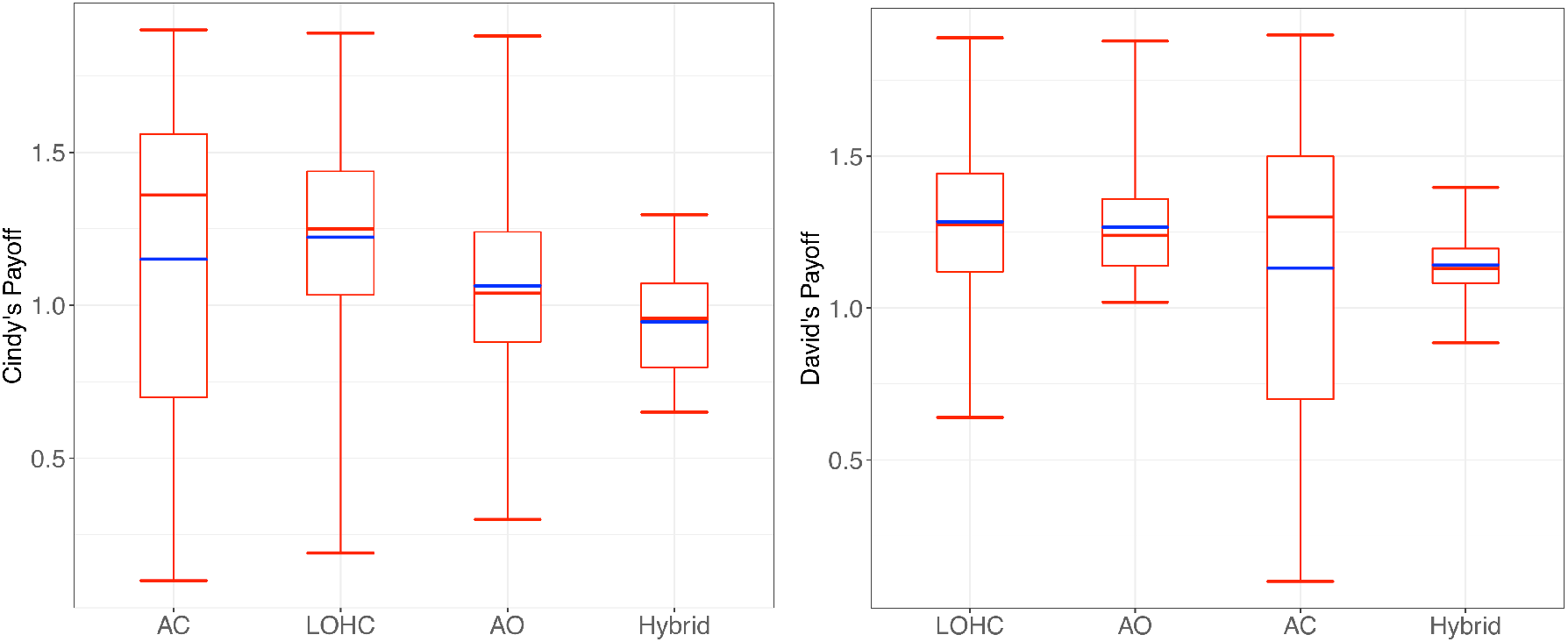
Cindy’s and David’s payoffs under different policies are illustrated as standard box charts respectively. Notably, the order of payoffs is different for Cindy (the left chart) and David (the right). With Cindy’s payoff, the descending order (from left to right) is *AC>LOHC>AO>Hybrid*, and with David’s payoff, the descending order is *LOHC>AO>AC>Hybrid*. The orders are statistically significant per Wilcoxon tests (*P*-value<0.05). Note the scatter points are omitted in the box charts to avoid overwhelmingly large number of points, which could lead to overly big file sizes of the graph.

## Conclusions and Discussion

### Conclusions

The main insights from the CDC game can be summarized as follows:

i. Among the 16 (4×4) equivalently reciprocal strategy interactions of the CDC game, approximately ½ are unnatural or irrational, which allows us to capture not only appropriate strategies but also those unnatural (or irrational) strategies that might be generated by faulty judgment(s) and/or miscommunications in policy-making system. An advantage of our CDC game approach is the completeness of the 16-elements strategy set in a two-player game.
ii. The CDC game has six types of equilibriums, which are evolutionarily stable and can be sustainable in practice. Some of the equilibriums are Nash equilibrium, which means that no game player would regret for his or her strategy. Some are pooling equilibrium, which can be roughly considered as “dictatorship-style”; in the case of CDC game, the pooling equilibrium implies that David would ignore Cindy’s advice. Some are separating equilibrium, which can be roughly considered as the opposite to pooling equilibrium, *i.e*., Cindy’s advice does make differences. The so-termed polymorphic hybrid equilibrium (PHE) means that sometimes the equilibrium is pooling and sometimes separating.
iii. There are four basic anti-pandemic policies: LOHC, HCLO, AO, and AC. They can be mapped to the output of CDC game system, *i.e*., the ultimate decision made by David in the CDC game. The LOHC is arguably the most rational policy and has been adopted by the majority of countries (regions) in the world. The diametrically opposite HOLC is obviously the least rational, and it is unlikely being adopted unless fatal mistake occurs. Although the HOLC can be mapped to strategy interactions ***B*** & ***E***, both do not have any equilibriums, mirroring the reality that the policy is unlikely to sustain in practice. In contrast, the LOHC policy can be mapped to strategy interactions, ***A, F***, and ***PHS*** (polymorphic hybrid strategy), with ***A*** being Nash equilibrium, ***F*** being ESS (evolutionarily stable strategy), and ***PHS*** being hybrid (sometimes pooling and sometimes separating). The simulation (of 100-millions times) with the CDC game revealed that the probability of LOHC is small-probability event (*P*=0.053), while the probability of HOLC is zero. The small-probability event characteristic of LOHC appears to be counter-intuitive, and we postulate that practical implementation of LOHC can be too complex to be feasible, especially for the COVD-19 pandemic that has been recurring one wave after another. The multi-wave and extreme contagiousness of COVID make failure in one round of reopening sufficiently ruin the whole efforts, *i.e*., cascading failures.
iv. The AO policy can be mapped to the equilibriums of mixed strategy of *J* & *K*, or PHS. The mixed strategy *J* & *K* are pooling equilibrium, which means that the CDC chief scientist’s advice makes no difference or ignored by the ultimate decision-maker, David. The hybrid strategy PHS may also generate AO policy, and in this case, sometimes the chief’s advice matters and sometimes does not. The simulation of 100-million times with the CDC game revealed that the probability of AO policy is *P*=0.412, and that of PHS is negligible (*P*=0.002). Therefore, virtually all AO policies are generated from the mixed-strategy of *J* & *K*, which is from David’s “dictatorship-style” decision (a characteristic of pooling equilibrium).
v. The AC policy can be mapped to the equilibriums of mixed strategy *I* & *L*, or PHS. Simulations revealed that the probability of AO policy is *P*=0.533, and that of PHS is negligible (*P*=0.002). Similar to previous AO policy, the equilibriums of *I* & *L* are pooling, and therefore, Cindy’s advice usually does not matter in the making of AC policy.
vi. Simulations of 100 million times suggested that the order of probabilities of three policies is AC>AO>LOHC, while HOLC=0. Interestingly, in terms of the payoff, Cindy and David have different orders. For Cindy, her payoff order is AC>LOHC>AO; for David, his payoff order is LOHC>AO>AC. Note that these three orders are of different nature. The first order is the likelihood of policy order, which is the product of CDC game, or the outcome of Cindy and David strategic interactions. The second order is Cindy’s preference (payoff) order, which should largely depend on her professional expertise; her profession as CDC chief obviously should have pushed her to prefer AC, *i.e*., her payoff (such as reputation or satisfaction from saving lives) is maximized. The third order is David’s preference (payoff) order, which should largely depend on his duty as president who needs to balance health *vs*. wealth. For president, AC is likely his last favorite, while LOHC is ideally his top choice although it is often too complex to implement. Therefore, although both Cindy and David seem to have different preferred strategies, their interactions produce a single set of “meta-strategies”, *i.e*., the policy set of LOHC, AO, and AC.
vii. More or less, most countries (regions) in the world seemed to have adopted the LOHC policy, while a handful of countries have adopted *laissez-faire* strategy such as passive herd immunity that may be considered as AO policy. The dynamic zero-out policy adopted by several countries (notably China and New Zealand) seemed to be equivalent to AC in terms of the effects, and to “precision” LOHC operationally in terms of the implementation.

In conclusion, through rigorous analytic and extensive simulation analyses of the CDC game, we obtained reasonable interpretations for the three fundamental public-health policies against the COVID-19 pandemic, *i.e*., AO, AC and their middle ground LOHC, precluding the obviously irrational or anti-intelligent HOLC policy. The policy of AO is likely to be wealth-optimal, while the policy of AC is likely to be health-optimal. The LOHC may achieve some level of balance between health and wealth, given it is a middle ground of both AC and AO. The HOLC generally favors neither health nor wealth, what we called anti-intelligent and is not sustainable (corresponding to no equilibriums in the CDC game). In real world, most countries (regions) have adopted the LOHC policy, which seems to balance the health *vs*. wealth dilemma and appears to be an optimal policy choice. However, simulations suggest that the policy may be too complex to implement successfully given its probability is only 0.053, a typical small probability event. In contrast, the two ends of the policy spectrum, AO and AC exhibited large probabilities (close to 0.41∼0.53). A take-home message seems to be KISS (keep it simple strategically and persistently), and the large probabilities of AC and AO are likely due to their simplicity.

### Discussion

Humans are believed to possess exceptional evolutionary capacity for decision-making, which should have contributed to the dominance of the humans on the earth planet (Samson 2020). Although on macroscopic scale, humans have been enormously successful from selecting the right food and shelter, to devising complex economic strategies and effective public health policies (Samson 2020), we have occasionally made expensive and painful mistakes, locally, regionally and globally, including the strategies and policies for fighting COVID-19 pandemic. The challenge seems particularly serious in the era of disinformation (*e.g*., Bergstrom & West 2020). Given the nature of the complexity and challenges, we formulate the decision-making problem for anti-COVID-pandemic as the CDC game, which is an extension to the classic SPS game. The latter achieved wide success in studying the reliability (honesty) of animal communications in evolutionary biology. The SPS game, especially its extensions (Huttegger & Zollman 2010, Whitmeyer 2020, Ma & Zhang 2021) are particularly powerful in modeling the processes that are driven by 3C forces (cooperation, competition and communication). Since we are only interested in strategic decision-making, it is natural for us to focus on the stable equilibriums. Tactical or operational level analyses are not considered in this article.

There are 16(4×4) possible strategy combinations (interactions) with the CDC game given it is an asymmetric four-by-four strategic-form game. Also given the completeness of the strategy interactions (4×4=16), the strategy interactions are in effects *reciprocal*. That is, there is likely an irrational (or unnatural) counterpart for each rational (or natural) strategy interaction. Given the equivalently reciprocal nature of the CDC game, it is expected that approximately ½ of the strategy interactions can be unnatural. Furthermore, there are *six* types of *equilibriums* (Table 2) among the 16 possible strategic interactions (Table 1 and Table 2). The *six equilibriums* can be classified as separating (signaling), pooling, and polymorphic hybrid equilibriums, and they may possess different behavioral and payoff properties (*see* Table 3 and Box S1 & S2). These equilibriums and stabilities can be significantly different, with different theoretical ramifications (*e.g*., Maynard-Smith & Harper 2003, Huttegger & Zollman 2001, Bergstrom & Lachmann 1997, 1998, Biernaskie *et al* 2018, Madgwick & Wolf 2020, Whitmeyer 2020). Practically, these six equilibriums can be mapped to three basic anti-pandemic policies.

Among the four fundamental anti-pandemic policies, the LOHC (low-risk open and high-risk closed) is essentially a middle ground of the two extreme policies: AC (always closed) and AO (always open). The LOHC policy appears to be the most rational and apparently most effective, and it has obviously been adopted by most countries (regions) of the world for fighting against COVID19 pandemic. Herd immunity or co-existence policies can be considered as AO policies. The strict AC policy, although medically optimum but economically risky, seems to be the least popular. In summary, co-existence with virus *vs*. zero-out virus appears to mirror the AO *vs*. AC policies, while LOHC policy seems to be the middle ground of both AO and AC. The unnatural HOLC (high-open and low-closed) is irrational, and have been adopted by nobody in the world. Strategy interactions B & E can be mapped to the HOLC policy, but they do not have any equilibriums, which is expected and simply mirrors the unsustainable nature of the HOLC policy.

Among the 16 (4×4) strategy interactions, approximately ½ of which can produce equilibriums, which can be mapped to the three fundamental policies as discussed previously. For the approximately ½ of the strategy interactions that do not have corresponding equilibriums, their strategies are unsustainable in practice and may mirror some of the unnatural and idiosyncratic behaviors in anti-pandemic policy-making. The explanation for the approximately ½ natural *vs*. unnatural strategies or with/without corresponding equilibriums should be to do with the reciprocal nature of the strategy set (options) as explained previously. Although it is difficult to interpret all of the unnaturalness or the idiosyncrasy in half of the strategy set, there are some answers from the recent studies in behavior economics (Samson 2014, 2020). Studies have revealed that humans are not always self-interested, benefits maximizing, and costs minimizing with stable preferences. Human minds are not always rational and may only possess *bounded rationality*, which suggests that human rationality is limited by brain’s information processing capability, insufficient knowledge feedback, and time constraint (Ma 2015a, 2015b, Samson 2014, 2020), not to mention complex politics in the case of CDC game. The fact that approximately ½ of the possible strategy options seem to demonstrate the rather high likelihood to devise an unsustainable (faulty) policy. In other words, behaving rationally should not be considered as granted for the CDC-president interactions.

In contrast, the approximately ½ of the strategy interactions with possible equilibriums can be sustainable, and mapped to one of the three basic policies (AO, AC, & LOHC). However, equilibriums, stability, and rationality do not necessarily correspond to scientifically optimal, and may not even necessarily correspond to scientific correctness, since the CDC output (policies) may also be influenced by politics or other socioeconomic factors. This is determined by the aim of this study—identifying the potential cognitive gaps and possibly traps by emulating the reality of anti-pandemic policies with the CDC game model.

The AO policy is essentially the *laissez-faire* strategy (*e.g*., passive herd immunity), which was initially adopted by a handful of countries including Sweden and partially by UK. The strictly AC seems unrealistic intuitively, but theoretically it does corresponds to two strategies with stable equilibriums. In practice, the dynamic zero-out (infections) policy adopted in China and New Zealand can be considered as either precision version of LOHC in terms of implementation, or an equivalent AC policy in terms of the effects. The dynamic zero-out policy does have a precondition, namely, it must be feasible to control the infection level below the threshold of *Allee effect*, and therefore, the infections level should be relatively lower in general to further push it below the threshold (of “local extinction”).

The Allee effect is named after animal ecologist W. C Allee (Allee & Bowen 1932), and it is a theoretic threshold of population growth (such as the infections of COVID-19), above which population growth may accelerate and below which population growth may decelerate and may go extinct ultimately (Kramer *et al*. 2017). Its implications to epidemiology have been discussed in recent literature; for example, Friedman *et al*. (2012) studied the relationship between the Allee effect of disease pathogens and the Allee effect of hosts (*e.g*., humans) in order to answer a question of fundamental importance in epidemiology: Can a small number of infected individual hosts with a fatal disease drive the host population to go extinct (assuming a healthy stable host population at the disease-free equilibrium is subject to the Allee effect)? Ma (2020) estimated the population aggregation critical density, the threshold for aggregated (clustered) infections of COVID-19 to occur. Besides Allee effects, two other theories, *i.e*., metapopulation dynamics and tipping-point theories, are particularly important for supporting the dynamic zero-out policy. The metapopulation theory maintains that extinctions of local population are common events, and tipping point theory suggests that, at some critical points, population growth may transit to either outbreak or die-off from the tipping point (threshold) (Citron *et al*. 2021). There are techniques for detecting tipping points by identifying some early warning signals (EWS), which can help to implement dynamic zero-out policy. These theories suggest that there are thresholds (tipping points) at which local pathogen (infections) may go extinct, depending on the dynamics of host-pathogen system and disease control measures. Therefore, public health policies designed to drive the infections (such as SARS-COV-2) to go extinct *locally* (below the threshold of Allee effect of the pathogen), while keep the host population absolutely safe from the risk associated with the Allee effect of the hosts, should be feasible. In other words, the dynamic zero-out policy, which is equivalent to driving *local* infections to extinctions, is theoretically feasible when implemented properly.

## Data Availability

All data generated from this study are from computer simulation and included in the Supplementary Tables S1-S4. No data related to human subjects (neither biometric data nor their behavioral measures) are used in this study and no ethical approval is applicable. Supplementary Tables S1-S4 are available upon request, and not provided here due to file-size limitation.

## Acknowledgements

This study received funding from the Cloud-Ridge Industry Technology Leader Grant of Yunan Province. We are indebted to Daisy Chen, from the Chinese Academy of Sciences, for her computational assistance.

## Online Supplementary Information for

### Box S1.

Three Key Elements (*Who, What* and *How*) to Formulate CDC Games and the Exposition of the Transformation from SPS through to CDC game, and CDC Parameters

Online Supplementary Information for “CDC (Cindy and David’s Conversations): Advising President to Survive Pandemic”
| **Box S1. Three Key Elements (*Who*, *What* and *How*) to Formulate CDC Games and the Exposition of the Transformation from SPS through to CDC game, and CDC Parameters** |  |
| --- | --- |
| (i) *Who* are the game players | **Sender** (Cindy or Adviser) *vs.* **Responder** (David or Decision-maker). **Sender** (signaler) was Sir Philip’s comrade, and **Responder** was Sir Philip Sydney in Maynard-Smith (1991) original SPS game. In the CDC game, Cindy is the message *sender* or *adviser*, and David is the *responder* or *decision-maker*. The essentiality of the role assignment lies in that who signals and who responds. |
| (ii) *What* is the content of message? | It is apparently the “request for water bottle” for the message **sender**, and the **responder** could either honor or ignore the request (water bottle) in the classic SPS game. The essentiality here is the message of “state-of-the-risk” (injured in SPS or high-risk in CDC) with probability of *m*. |
| (iii) *How* is message sent (signaled)? | Through shouting or whispering in classic SPS game. In the CDC game, it is through some communication channel such as phone, person conversation, or media. However, the ‘*How*’ point does not matter much in the sense that it does not influence the *mathematical logic* of the game. |
| 4 (2x2)—Strategy Set in Classic SPS Game | There were only *four* (2x2) strategies in classic (Maynard-Smith 1991) SPS game since it was considered sufficient for its initial design, *i.e.*, to demonstrate the *handicap* principle or the *reliability* (*honesty*) of animal communications (Zahavi 1975, 1997). The initial hypothesis of the handicap principle was that to communicate honestly, there must be a handicap in the communication system that enforces the honesty. With the handicap principle, only four strategies that make sense. We use the term “make sense” to imply that the strategies are broader than being “rational” since evolutionary game theory does not enforce “rationality.” Instead, as long as there are ESS (evolutionary stable strategy) equilibriums, the strategies make sense. Actually, the strategies in the original SPS game not only sensible (natural), but also are rational, including sending dishonest signal (selfish behavior) since payoff was high for doing so. |
| 16(4x4)-Strategy Set in the CDC Game | It was Huttegger & Zollman (2010), Whitmeyer (2020) and *etc.*, which significantly expanded the classic SPS game, most notably, expanding the strategies from 4 to 16, which transformed the asymmetric, classic SPS to an *equivalently* reciprocal game given virtually all possible strategies are allowed. This extension allowed Ma & Zhang (2021) to describe various idiosyncrasies in masking behaviors in the context of ABD (Alice and Bob’s dating dilemma) game.<br>Table 1 and Fig 1 listed the 16 possible strategies of the CDC game. The *strategy matrix* starts with (C<sub>1</sub>, D<sub>1</sub>) strategy: Open only if low-risk; D<sub>1</sub>: Closed only if Cindy advises closed. This is arguably one of the most rational “conversations” (advice-decision) in the CDC game. Once the first strategy pair (C<sub>1</sub>, D<sub>1</sub>) is specified, the remaining 15 pairs are set to keep consistent mathematic logics, as well as reciprocal interactions. The logic consistency refers to the validity of mathematic logic, which determines the rigorous inferences of the various equilibriums listed in Table 2. The equilibriums indicate that the corresponding strategies can be evolved and become established (adopted) in a society. Nevertheless, the validity of the mathematic logic does not guarantee the rationality or naturalness of strategies. That is, valid strategies can be senseless, mirroring various idiosyncratic decisions during the pandemic. |
| Potentially Alternative Strategy Matrices (Sets) for CDC game | Before finalizing the CDC strategy matrix as exhibited by Table 1 & Fig 1, we tried several alternatives. We realized that, after elements (*i: Who*) & (*ii: What*) are specified, the setting of (*iii: How*) does not matter in the sense that the setting only affects the *order* of the 16 combinatorial (interaction) strategies. It influences neither the validity of various strategy *equilibriums* (Table 2) nor the *completeness* (Table 1 or Fig 1) of the strategy set (options). That is, whatever order is specified, all possible strategy options (behaviors) are covered by the game (*i.e.*, completeness). That is, although the classic SPS (with 4 strategy pairs) is not reciprocal, in the extended SPS game including CDC game, when the 12 ignored strategy pairs (due to the non-reciprocal or asymmetrical nature of SPS) were brought back. In effects, the CDC games cover all possible strategy options (*completeness*), and the strategic interactions are reciprocal due to the completeness. |
| **Interpretations of the parameters (*a*, *b*, *c*, *d*, *m* & *k*) in the SPS/CDC Games** |  |
| *a*, *b*, *c*, *d* | Parameters *a* and *b* the cost parameters associated with Cindy, with *a* is the cost of misjudgment of high risk (underestimate risk), and *b* of low risk (overestimate risk), respectively. Parameter *c* is the strategic cost from the mistake in the CDC system. Parameter *d* is the cost parameter associated with David’s decision-making such as the decline of public support. |
| *m* & *k* | Parameter *m* and *k* are different from the previous previously defined parameters *a*, *b*, *c*, and *d* in the sense that they may influence ‘cost’, but they are not ‘cost’ *per se*. Parameter *m* is the risk level Cindy judges in a country (region), and parameter *k* is the relatedness between Cindy and David. Parameter *k* represents their shared interests (shared genes in evolutionary biology) and it has a |
|  | direct impact on their *inclusive fitness (payoff)* (=k times the fitness of the other player plus his or her own fitness). |

### Box S2.

Definitions for the Equilibrium Types and Stability Properties in the SPS/CDC Games

| **Box S2. Definitions for the Equilibrium Types and Stability Properties in the SPS/CDC Games** |  |
| --- | --- |
| **Definitions for the Equilibrium Types** |  |
| **Nash equilibrium** | A *Nash equilibrium* (i.e., “no-regret” strategies) is a solution to a non-cooperative game involving two or more players, in which each player is supposed to be aware of the equilibrium strategies of the other players, and no player is better off by unilaterally changing own strategy. In other words, when each player has selected a strategy and no player may benefit from changing strategies whereas the other players keep theirs fixed, then the current set of strategies and the consequent payoffs (fitness) constitute a Nash equilibrium. |
| **ESS (evolutionarily stable strategy)** | An *evolutionarily stable strategy* (ESS) is a strategy that, if adopted by a population of players in a given environment, cannot be invaded by any mutational (alternative) strategy, which is usually scarce initially. It is a Nash equilibrium that is “evolutionarily stable or resilient.” In other words, an ESS is a strong version of the Nash equilibrium, and the Nash equilibrium is a weak version of ESS. |
| **Pooling equilibrium** | A *pooling equilibrium* is an equilibrium in which, regardless of the types of sender with differing characteristic signals, the receiver shall choose the same response. In the case of the CDC game, David does not care whether what Cindy advises when he makes the decision. |
| **Separating equilibrium** | A *separating equilibrium* is opposite to pooling equilibrium: a receiver’s response can be different depending on the type of sender’s signal. In the case of CDC game, Cindy’s advice does influence David’s decision. |
| **Hybrid equilibrium** | A *hybrid equilibrium* is some combination of *separating* equilibrium and *pooling* equilibrium. |
| **Polymorphism** | A *polymorphism* is a mixed Nash equilibrium where sender mixes between being honest and dishonest signaling, and responder may also adopt a mixed strategy accordingly. Polymorphisms, which were ignored in biological signaling until Huttegger & Zollman (2010) extensions of the original SPS game, correspond to hybrid equilibria in this study. Polymorphic equilibrium allows for partially meaningful (honest) communication, and therefore grants the SPS much realism. |
| **Definitions for the Stability Properties** |  |
| **Lyapunov Stable** | Assume a dynamical system which is described with<br>$\dot{x} = f(x), \quad x \in R^n,$<br>there is an equilibrium solution $\bar{x} \in R^n$, such that $f(\bar{x}) = 0$. The equilibrium solution $\bar{x}$ is stable in the sense of Lyapunov stable, if for given $\varepsilon > 0$, there exists $\delta = \delta(\varepsilon) > 0$ such that, for any other solution $y(t)$, $\ y(t_0) - \bar{x}\ < \delta$ ($t_0 \in R$), then $\ y(t) - \bar{x}\ < \varepsilon$ for $t > t_0$.<br>Lyapunov stability is a rather weak requirement on equilibrium points. Especially, it does not require that trajectories starting close to the origin approach to the origin asymptotically. In a simplified interpretation, if the solutions that start out near an equilibrium point $x$ forever, then $x$ is Lyapunov stable. |
| **Asymptotically Stable** | A time-invariant system is asymptotically stable if all the eigen-values of its system matrix (**A**) possess negative real parts. In the case of asymptotic stability, there is a sphere $S$, centered around $\delta = 0$ with the radius $r$, such that the response, once entered the sphere, converges to the origin. |
| **Neutrally Stable** | An equilibrium that is Lyapunov stable but not asymptotically stable is sometimes termed as *neutrally stable*. |

## References

Allee, WC, Bowen E (1932) Studies in animal aggregations: mass protection against colloidal silver among goldfishes. Journal of Experimental Zoology. Vol. 61(2):185–207.

Amadae, S (2016) Prisoner’s Dilemma, Prisoners of Reason. Cambridge University Press, NY, pp. 24–61.

Aumann, R (1959) Acceptable points in general cooperative n-person games. In Luce, R. & Tucker, A W (eds.) Contributions to the Theory of Games IV. Annals of Mathematics Study. Vol. 40. Princeton University Press.

Axelrod, R & Hamilton, WD (1981) The evolution of cooperation. Science, 211, 1390–1396.

Bergstrom, C. T. & Lachmann, M (1997) Signalling among relatives I. Is costly signaling too costly? Phil. Trans. R. Soc. Lond. B 352, 609–617.

Bergstrom, C. T. & Lachmann, M (1998) Signalling among relatives III. Talk is cheap. Proc. Natl Acad. Sci. USA 95, 5100–5105.

Bergstrom, C. T. and J. D West (2020) Calling Bullshit: The Art of Skepticism in a Data-Driven World. Allen Lane.

Biernaskie, JM, JC Perry, and A Grafen (2018) A general model of biological signals, from cues to handicaps. Evolution Letters 2-3: 201–209

Brown, JS. 2016 Why Darwin would have loved evolutionary game theory. Proc. R. Soc. B, 283: 20160847. http://dx.doi.org/10.1098/rspb.2016.0847

Citron, DT, CA Guerra, AJ Dolgert, SL Wu, JM Henry, and DL Smith (2021) Comparing metapopulation dynamics of infectious diseases under different models of human movement. PNAS, Vol. 118 No. 18 e2007488118

Cooper, GA, SR Levin, G Wild, & SA West (2018) Modeling relatedness and demography in social evolution. Evolution Letters 2-4: 260–271

Friedman, A & AA Yakubu (2012) Fatal disease and demographic Allee effect: population persistence and extinction, Journal of Biological Dynamics, 6:2, 495–508

Grafen, A. (1990) Biological signals as handicaps. J. Theor. Biol. 144, 517–546.

Hilker, F, Langlais, M & Malchow, H (2009) The Allee Effect and Infectious Diseases: Extinction, Multistability, and the Disappearance of Oscillations. The American Naturalist. Vol. 173. 72–88.

Hamilton, WD (1964) The genetic evolution of social behavior. Journal of Theoretical Biology. vol. 7:1–32.

Huttegger, S. & KJS Zollman (2010) Dynamic stability and basins of attraction in the Sir Philip Sidney game. Proc. R. Soc. B. vol. 277, 1915–1922.

Jones, DS (2020) History in a Crisis—Lessons for Covid-19. New Engl. J. Med., Vol. 382;18, http://nejm.org April 30, 2020

Kramer, AM, L Berec, JM Drake (2017) Editorial: Allee effects in ecology and evolution. vol. 87(1):7–10, https://doi.org/10.1111/1365-2656.12777

Levins, R (1969) Some demographic and genetic consequences of environmental heterogeneity for biological control. Bulletin of the Entomological Society of America, 15 (3): 237–240

Ma ZS (2009) Towards an extended evolutionary game theory with survival analysis and agreement algorithms for modeling uncertainty, vulnerability, and deception. Springer Lecture Notes in Artificial Intelligence, vol. 5855: 608–618.

Ma ZS, AW Krings, and FT Sheldon (2010) The handicap principle, strategic information warfare and the paradox of asymmetry. The 6th Cyberspace Sciences and Information Intelligence Research Workshop, 6th CSIIRW10, Oak Ridge National Lab, Oak Ridge, USA.

Ma ZS & Krings AW (2011) Dynamic hybrid fault modeling and extended Evolutionary Game theory for reliability, survivability and fault tolerance analyses. IEEE Transactions on Reliability, 60(1):180–196.

Ma ZS (2015a) Towards computational models of animal cognition, an introduction for computer scientists. Cognitive Systems Research, 33:42–69.

Ma ZS (2015b) Towards computational models of animal communication, an introduction for computer scientists. Cognitive Systems Research, 33:70–99.

Ma ZS (2020) Predicting the Outbreak Risks and Inflection Points of COVID-19 Pandemic with Classic Ecological Theories. Advanced Science, https://doi.org/10.1002/advs.202001530

Ma ZS & YP Zhang (2021) To mask, or not to mask, Alice and Bob’s dating dilemma. (Under Review)

Madgwick, P. & JB Wolf (2020) Evolution of strategic cooperation. Evolution Letters 4-2: 164–175

Maynard Smith, J. & Price, G. (1973) The logic of animal conflict. Nature 146, 15–18.

Maynard Smith, J (1991) Honest signaling: the Philip Sidney game. Animal Behavior, 42:1034–35.

Maynard Smith, J. & Harper, D. (2003) Animal Signals. Oxford, UK: Oxford University Press.

Nason Maani & Sandro Galea (2021) https://www.scientificamerican.com/article/what-science-can-and-cannot-do-in-a-time-of-pandemic/

Nicholas Dirks (2021) https://www.scientificamerican.com/article/we-need-social-science-not-just-medical-science-to-beat-the-pandemic/

Nowak MA. 2006 Five rules for the evolution of cooperation. Science 314, 1560–1563.

Nowak MA & K Sigmund (2007) How Populations Cohere: Five Rules for Cooperation, in “Theoretical Ecology: Principles and Applications”3rd Edited by Robert May & Angela McLean

Rasmussen, S. & DJ Jamieson (2020) Public Health Decision Making during Covid-19—Fulfilling the CDC Pledge to the American People. New Engl. J. Med. vol. 383;10 http://nejm.org, September 3, 2020.

Samson, A. (Ed.)(2014). The Behavioral Economics Guide 2014 (with a foreword by George Loewenstein and Rory Sutherland) (1st ed.). Retrieved from http://www.behavioraleconomics.com

Samson, A. (Ed.)(2020). The Behavioral Economics Guide 2020 (with an Introduction by Colin Camerer) Retrieved from https://www.behavioraleconomics.com

Tanimoto J (2015) Fundamentals of Evolutionary Game Theory and its Applications. Springer.

Tanimoto J (2021) Sociophysics Approach to Epidemics. Springer.

Whitmeyer, M (2020) Strategic inattention in the Sir Philip Sidney game. bioRxiv preprint doi: https://doi.org/10.1101/559955

Zahavi, A (1975) Mate selection—a selection for a handicap. J. Theor. Biol. 53, 205–214.

Zahavi, A & A Zahavi (1997) The handicap principle: a missing piece of Darwin’s puzzle. Oxford University Press.

